# Brain atrophy in prodromal synucleinopathy is shaped by structural connectivity and gene expression

**DOI:** 10.1101/2021.12.27.21268442

**Authors:** Shady Rahayel, Christina Tremblay, Andrew Vo, Ying-Qiu Zheng, Stéphane Lehéricy, Isabelle Arnulf, Marie Vidailhet, Jean-Christophe Corvol, ICEBERG Study Group, Jean-François Gagnon, Ronald B. Postuma, Jacques Montplaisir, Simon Lewis, Elie Matar, Kaylena Ehgoetz Martens, Per Borghammer, Karoline Knudsen, Allan Hansen, Oury Monchi, Bratislav Misic, Alain Dagher

**Affiliations:** The Neuro (Montreal Neurological Institute-Hospital), McGill University, Montreal H3A 2B4, Canada; Centre for Advanced Research in Sleep Medicine, Hôpital du Sacré-Cœur de Montréal, Montreal H4J 1C5, Montreal, Canada; Wellcome Centre for Integrative Neuroimaging, Centre for Functional Magnetic Resonance Imaging of the Brain, University of Oxford, John Radcliffe Hospital, Oxford OX3 9DU, United Kingdom; Sorbonne Université, Institut du Cerveau – Paris Brain Institute – ICM, INSERM, CNRS, Assistance Publique Hôpitaux de Paris, Pitié-Salpêtrière Hospital, Paris 75013, France; Department of Psychology, Université du Québec à Montréal, Montreal H2X 3P2, Canada; Research Centre, Institut universitaire de gériatrie de Montréal, Montreal H3W 1W5, Canada; Department of Neurology, Montreal General Hospital, Montreal H3G 1A4, Canada 7 Department of Psychiatry, Université de Montréal, Montreal H3T 1J4, Canada; ForeFront Parkinson’s Disease Research Clinic, Brain and Mind Centre, University of Sydney, Camperdown NSW 2050, Australia; Department of Kinesiology and Health Sciences, University of Waterloo, Waterloo N2L 3G1, Canada; Department of Nuclear Medicine and PET, Aarhus University Hospital, Aarhus DK- 8200, Denmark; Departments of Clinical Neurosciences, Radiology, and Hotchkiss Brain Institute, University of Calgary, Calgary T2N 4N1, Canada

**Keywords:** REM sleep behaviour disorder, Parkinson’s disease, dementia with Lewy bodies, alpha-synuclein, MRI

## Abstract

Isolated REM sleep behaviour disorder (iRBD) is a synucleinopathy characterized by abnormal behaviours and vocalizations during REM sleep. Most iRBD patients develop dementia with Lewy bodies, Parkinson’s disease, or multiple system atrophy over time. Patients with iRBD exhibit brain atrophy patterns that are reminiscent of those observed in overt synucleinopathies. However, the mechanisms linking brain atrophy to the underlying alpha-synuclein pathophysiology are poorly understood. Our objective was to investigate how the prion-like and regional vulnerability hypotheses of alpha-synuclein might explain brain atrophy in iRBD.

Using a multicentric cohort of 182 polysomnography-confirmed iRBD patients who underwent T1-weighted MRI, we performed vertex-based cortical surface and deformation-based morphometry analyses to quantify brain atrophy in patients (67.8 years, 84% men) and 261 healthy controls (66.2 years, 75%) and investigated the morphological correlates of motor and cognitive functioning in iRBD. Next, we applied the agent-based Susceptible-Infected-Removed model (i.e., a computational model that simulates in silico the spread of pathologic alpha-synuclein based on structural connectivity and gene expression) and tested if it recreated atrophy in iRBD by statistically comparing simulated regional brain atrophy to the atrophy observed in patients. The impact of *SNCA* and *GBA* gene expression and brain connectivity was then evaluated by comparing the model fit to the one obtained in null models where either gene expression or connectivity was randomized.

The results showed that iRBD patients present with cortical thinning and tissue deformation, which correlated with motor and cognitive functioning. Next, we found that the atrophy simulated based on brain connectivity and gene expression recreated cortical thinning (*r*=0.51, *p*=0.0007) and tissue deformation (*r*=0.52, *p*=0.0005) in patients, and that the connectome’s architecture along with *SNCA* and *GBA* gene expression contributed to shaping atrophy in iRBD. We further demonstrated that the full agent-based model performed better than network measures or gene expression alone in recreating the atrophy pattern in iRBD.

In summary, atrophy in iRBD is extensive, correlates with motor and cognitive functioning, and can be recreated using the dynamics of agent-based modelling, structural connectivity, and gene expression. These findings support the concepts that both prion-like spread and regional susceptibility account for the atrophy observed in prodromal synucleinopathies. Therefore, the agent-based Susceptible-Infected-Removed model may be a useful tool for testing hypotheses underlying neurodegenerative diseases and new therapies aimed at slowing or stopping the spread of alpha-synuclein pathology.

## Introduction

Isolated REM sleep behaviour disorder (iRBD) is characterized by abnormal motor behaviours and vocalizations during REM sleep.^1, 2^ iRBD typically develops into dementia with Lewy bodies, Parkinson’s disease or multiple system atrophy,^3^ making it an early manifestation and phenotype of synucleinopathies. Specifically, iRBD is thought to result from the impairment of brainstem circuits involved in REM sleep muscle atonia that occurs as a result of early accumulation of pathologic alpha-synuclein in the pontine tegmentum.^4, 5^ However, MRI studies in patients with polysomnography-proven iRBD without cognitive or motor diagnoses have also shown patterns of diffuse brain atrophy reminiscent of dementia with Lewy bodies or Parkinson’s disease.^6, 7^ Moreover, the severity of cortical atrophy is a predictor of subsequent dementia, hence more severe disease.^8^

Dementia with Lewy bodies and Parkinson’s disease are thought to arise from the accumulation of misfolded alpha-synuclein in the brain.^9^ Previous pathological brain staging schemes have suggested that the toxic agent starts in the brainstem and then spreads upwards,^10, 11^ giving rise to the clinical features that lead to a dementia- or a parkinsonism- first phenoconversion in iRBD patients. Evidence from animal models shows that pathologic alpha-synuclein can propagate and promote protein misfolding, supporting the prion-like model of alpha-synuclein.^4, 12–17^ In line with this, MRI studies performed in patients with Parkinson’s disease also show that atrophy patterns are shaped by connectivity.^18–20^

We previously modelled alpha-synuclein propagation using a Susceptible-Infected-Removed (SIR) model,^21^ a computational model based on an adaptation of epidemiological SIR models but applied to neurological diseases with the underlying hypothesis that alpha-synuclein propagation works like an infection in a population. In this model, every agent is an autonomous alpha-synuclein molecule that can exist in three states: Susceptible (normal), Infected (misfolded) or Removed (degraded). In its misfolded state, the agent becomes infective. Agents can also move between regions via neural connections. The model uses *SNCA* and *GBA* gene expression as measures of local alpha-synuclein concentration, and connectivity to determine to determine agent numbers and propagation. The model has predicted atrophy distribution in Parkinson’s disease patients and alpha-synuclein distribution in a mouse model.^21, 22^ Null models show that both connectivity and local alpha-synuclein concentration are important factors shaping the propagation of agents.^21, 22^ However, it remains unknown if these mechanisms also explain the atrophy seen in early synucleinopathies.

In this study, we compiled neuroimaging data from several centres to generate a map of brain atrophy in iRBD. We then used the agent-based SIR model to test if brain connectivity and *SNCA* and *GBA* gene expression explain brain atrophy patterns in iRBD. Vertex-based cortical surface and deformation-based morphometry analyses were performed in 182 polysomnography-confirmed iRBD patients and 261 healthy controls who underwent T1-weighted brain MRI to characterize atrophy and investigate the correlates of motor and cognitive functioning. We then tested whether brain connectivity and gene expression recreated atrophy by simulating atrophy with the SIR model and by statistically comparing the simulated pattern to the actual atrophy pattern found in iRBD. We next used null modelling to test if connectivity and gene expression were decisive in shaping the brain atrophy seen in iRBD. We hypothesized that the SIR model would recreate brain atrophy in iRBD and that both connectivity and gene expression would be significant determinants of atrophy.

## Materials and methods

### Participants

A total of 443 participants (182 iRBD patients and 261 controls) were recruited from five sites: 116 (59 patients) from the Movement Disorders clinic at the Hôpital de la Pitié-Salpêtrière (France), 83 (48 patients) from the Centre for Advanced Research on Sleep Medicine at the Hôpital du Sacré-Cœur de Montréal (Canada), 56 (30 patients) from the ForeFront Parkinson’s Disease Research Clinic at the University of Sydney (Australia), 38 (18 patients) from Aarhus University Hospital (Denmark), and 150 (27 patients) from the Parkinson’s Progression Markers Initiative baseline cohort.^23^ All iRBD patients had a polysomnography-confirmed diagnosis of iRBD and were free of parkinsonism and dementia at the clinical examination closest in time to MRI.^24, 25^ An overview of the study protocol along with a flowchart of the selected patients are presented in Figure 1 and the cohorts’ demographics are available in Supplementary Table 1. All participants were part of research protocols approved by local ethics committees and provided written informed consent. The current project was also approved by the Research Ethics Board of the McGill University Health Centre.

**Figure 1.**
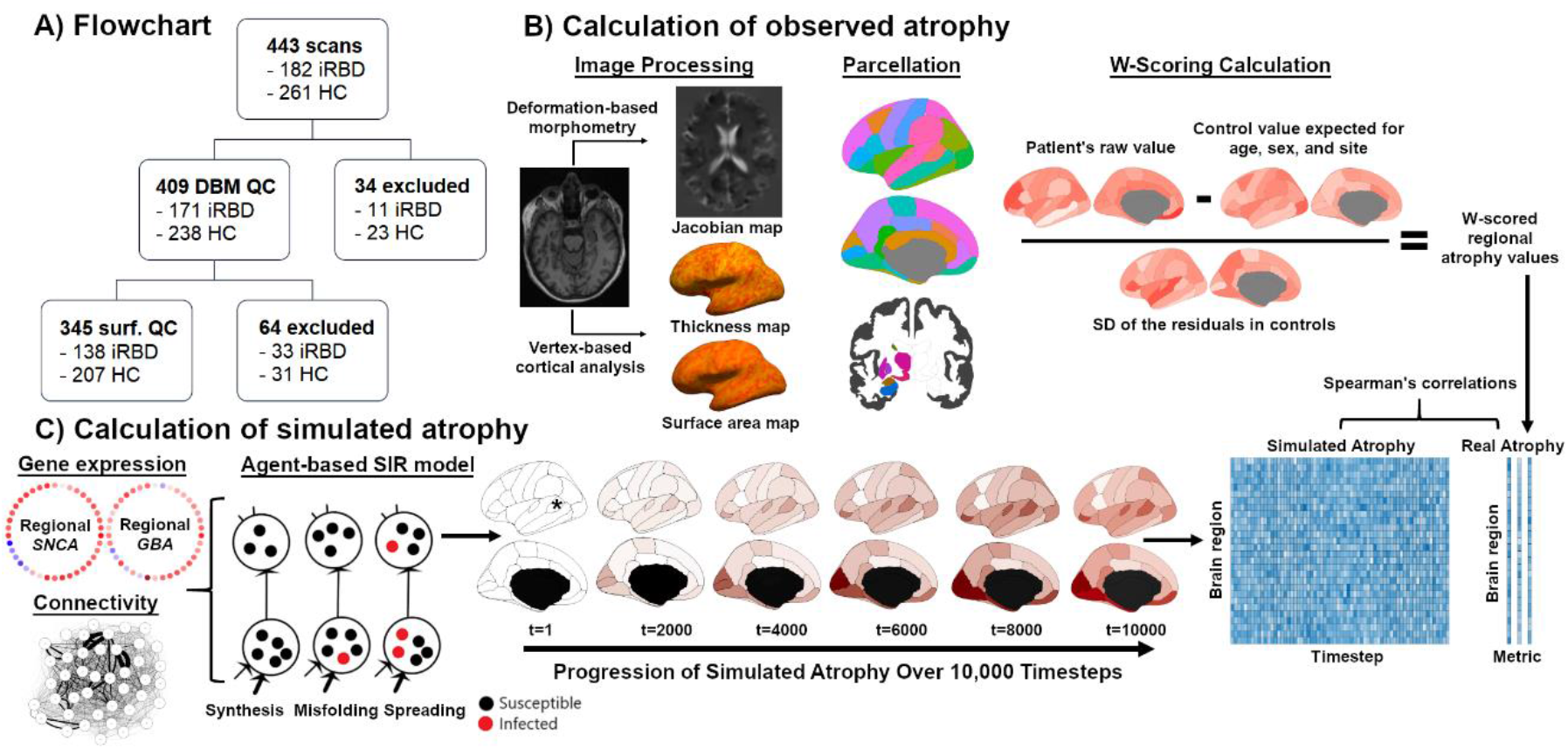
Demographics and clinical variables of iRBD and controls. **(A)** A flowchart of the different cohorts included in this study. **(B)** Overview of the study protocol for quantifying brain atrophy in iRBD. Deformation-based morphometry and vertex-based cortical surface analysis were performed to generate Jacobian and cortical thickness and surface maps. These were parcellated and *W*-scored to correct for the effects of age, sex, and site seen in controls. These regional values were the atrophy patterns to recreate. **(C)** The spread of alpha-synuclein in the brain was simulated in silico using the agent-based SIR model based on structural connectomics and *SNCA* and *GBA* gene expression. The simulation was iterated 10,000 times, with brain atrophy being simulated at each timestep. These patterns were correlated with the observed atrophy patterns to assess if the model recreated atrophy. HC = healthy controls; iRBD = isolated REM sleep behaviour disorder; QC = quality control; SD = standard deviation; SIR = Susceptible- Infected-Removed.

## MRI

### MRI acquisition

The Montreal cohort underwent T1-weighted imaging with a 3T Siemens TIM Trio scanner with a 12-channel head coil, MPRAGE sequence: TR: 2,300 ms, TE: 2.91 ms, flip angle: 9°, and voxel size: 1 mm³ isotropic. The Paris cohort underwent T1-weighted imaging with a 3T Siemens TIM Trio scanner with a 12-channel head coil, MPRAGE sequence: TR: 2,300 ms, TE: 4.18 ms, TI: 900 ms, flip angle: 9°, and voxel size: 1 mm³ isotropic; or a 3T PRISMA Fit scanner with a 64-channel head coil, MP2RAGE sequence: TR: 5,000 ms, TE: 2.98 ms, TI: 700 and 2,500 ms, flip angle: 4° and 5°, GRAPPA: 3, and voxel size: 1 mm³ isotropic. The Sydney cohort was imaged with a GE Discovery MR750 3T scanner with an 8-channel head coil, BRAVO sequence: TR: 5,800 ms, TE: 2.6 ms, flip angle: 12°, and voxel size: 1 mm³ isotropic. The Aarhus cohort was imaged with a 3T Siemens MAGNETOM Skyra scanner with a 32-channel head coil, MPRAGE sequence: TR: 2,420 ms, TE: 3.7 ms, TI: 960 ms, flip angle: 9°, and voxel size: 1 mm³ isotropic. The T1- weighted images from the Parkinson’s Progression Markers Initiative cohort, an international multicentre cohort, were also included (see www.ppmi-info.org for the imaging protocols).^23^

### Quantification of atrophy

Deformation-based morphometry was performed using CAT12 (version 12.7; www.neuro.uni-jena.de/cat) to quantify atrophy by measuring the non-linear change required in every voxel for registering the brain to the common template.^26^ The processing included bias correction, affine registration, unified segmentation,^27^ skull-stripping, parcellation, intensity transformation, partial volume estimation, and spatial normalization using DARTEL.^26^ This resulted in whole-brain maps of Jacobian determinants, which were smoothed with a 12-mm isotropic kernel and then used as the measure of local brain atrophy. Images with an automated quality rating below 80% were excluded from analyses involving deformation-based morphometry.

Brain atrophy in iRBD also manifests as abnormal cortical thickness and surface area.^28^ To ensure that findings were not due to the atrophy metric, the scans passing deformation-based morphometry quality control were also processed with FreeSurfer (version 6.0.0) to generate individual thickness and surface area maps of the whole cortex.^29, 30^ Every map was inspected by a trained rater (S.R.) and a score from 1-4 was assigned to each scan based on published guidelines;^31, 32^ scans with a score >2 (i.e., major reconstruction errors) were excluded from cortical surface analyses.

### *W*-scoring and brain parcellation

For deformation-based morphometry-derived atrophy, a *W*-score map was computed from each patient’s smoothed map by regressing out the effects of age, sex, and site found in the age- and sex-matched controls passing quality control.^33^ At each voxel, the following formula was applied:

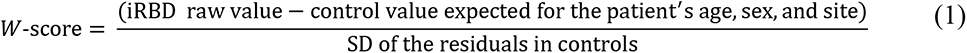

A voxel with a negative *W*-score represents greater atrophy (decreased volume) in the iRBD patient compared to controls, whereas a positive *W*-score indicates less atrophy (increased volume), while considering the confounds.

Regional *W*-scores were then extracted by parcellating every map with a 42-region atlas for which corresponding structural connectivity and gene expression data were available.^21^ This atlas comprised 34 cortical regions from the Desikan-Killiany atlas and 7 subcortical regions from FreeSurfer.^34, 35^ Due to its importance in synucleinopathies,^36, 37^ the substantia nigra was added from the 7-tesla “Atlas of the basal ganglia” (ATAG atlas).^38^ The 42 regional *W*-scores extracted from each map were then averaged across iRBD patients to yield 42 *W*-scores representing regional tissue deformation. *W*-scores were also extracted using finer parcellations of 65 and 119 regions to ensure that findings were robust to parcellation resolution. Because gene expression data, used by the model for determining regional alpha-synuclein synthesis and clearance (see below), were only available in the right hemisphere for 2 of the 6 existing post-mortem brains,^39^ and because diffusion tractography-based reconstruction of the connectome often leads to inaccurate measurements of interhemispheric connections,^40, 41^ the main analyses were performed in the left hemisphere.

For cortical surface analysis, thickness and surface area values were extracted from each surface from the same 34 cortical regions as used for deformation-based morphometry.^34^ Since subcortical regions do not have a cortical surface from which to derive thickness and area values, the global volume measurement generated as part of FreeSurfer’s subcortical processing was used. The substantia nigra was excluded from cortical surface analyses as it is unavailable in FreeSurfer, resulting in a total of 41 regions. Since cortical surface area and subcortical volume scale with head size,^42, 43^ the raw values were divided by the estimated total intracranial volume (derived from FreeSurfer). The same *W*-scoring procedure was then applied to these values to adjust for the effects of age, sex, and site from controls who passed quality control. To ascertain those findings were not due to the multicentric nature of the cohort, we also tested the model with harmonized regional *W*- scores derived from the ComBat harmonization method,^44–46^ a batch-effect correction tool used in genomics and validated for neuroimaging that removes the unwanted scanning-related inter-site variability while preserving biological variability.

Importantly, to facilitate interpretation when testing the model, the *W*-scores were inverted such that a positive score indicated a region with greater atrophy and a negative score indicated lower atrophy in iRBD patients compared to controls.

### Agent-based SIR model

#### Model overview

The agent-based SIR model (https://github.com/yingqiuz/SIR_simulator) was used to simulate the spread of alpha-synuclein.^21^ This algorithm applies agent-based modelling inside an SIR framework to model the spread as an epidemic shaped by the simultaneous effect of brain connectivity along with *SNCA* and *GBA* expression.^21^ This model previously recreated the atrophy observed in Parkinson’s disease and frontotemporal dementia.^21, 47^ In this model, every agent is an autonomous alpha-synuclein molecule that belongs to one of three mutually exclusive states: the “Susceptible” state when the agent is normal (normal alpha-synuclein), the “Infected” state when the agent becomes infective (misfolded alpha-synuclein isoform), and the “Removed” state when the agent is degraded. Agents can also move between regions via neural connections. The transitions between states are determined by rules guiding the interaction dynamics between agents and their regional environment. The model simulates atrophy in every brain region based on the following computational steps: 1) the production of normal alpha-synuclein, 2) the clearance of normal and misfolded alpha-synuclein, 3) the misfolding of normal alpha-synuclein, 4) the propagation of normal and misfolded alpha-synuclein, and 5) the emergence of atrophy.

#### Connectivity and gene data

The details of diffusion-weighted image processing, deterministic fibre tracking, and gene expression data have been described elsewhere.^21^ Briefly, the structural connectivity data were used to model the spread of agents from one region to another; they were derived from the pre-processed diffusion-weighted images of 1,027 participants from the Human Connectome Project.^61^ Deterministic tractography was used to construct consensus connectivity matrices between the 42 regions as described previously.^48–50^ The analysis was performed at connection densities of 25%, 30%, 35%, and 40% to ensure that results were robust to change in this parameter. A distance matrix of the mean Euclidean length of the corresponding streamlines for the 42 regions was also generated to modulate the rate of movement of agents between connected regions. For the gene expression data, the regional expression of *SNCA* and *GBA* were used to model the regional synthesis and clearance of alpha-synuclein; values were derived from the post-mortem mRNA transcription profiles of 6 subjects from the Allen Human Brain Atlas^39^ using *abagen* (https://abagen.readthedocs.io/en/stable/)51 and averaged for the 42 regions.

#### Production of normal alpha-synuclein (module 1)

In region *i*, the synthesis of susceptible agents per unit time occurs with probability *αi*:

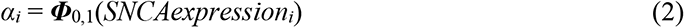

where ***Φ***_0,1_(·) is the standard normal cumulative distribution function and *SNCAexpression_i_* is the gene expression of *SNCA* of region *i*. At each timestep, the increment of susceptible agents in region *i* is *α_i_S_i_Δt*, where *S_i_* is the size of region *i* and *Δt* is the total time.

#### Clearance of normal and misfolded alpha-synuclein (module 2)

In region *i*, the clearance of susceptible and infected agents per unit time occurs with probability *βi*:

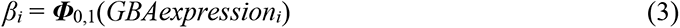

where *GBAexpression_i_* is the gene expression of *GBA* of region *i*. Considering that the probability that an agent is still active after *Δt* is given by lim*_δτ_*_→0_(1 − *βδτ*)^Δ*t*/*δτ*^ = *e*^−*β*Δ*τ*^, the cleared proportion within *Δt* is 1 – *e*^−*β*Δ*τ*^.

#### Misfolding of normal alpha-synuclein (module 3)

The susceptible agents not cleared from region *i* may become infected per unit time with probability *γ_i_*:

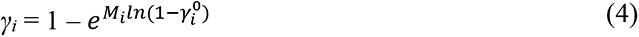

where *M_i_* is the population of infected agents in region *i* and 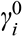 is the baseline likelihood that a susceptible agent becomes an infected agent in region *i*, which was set as the reciprocal of region size. The probability that a susceptible agent did not get infected is given by 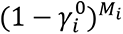; therefore, 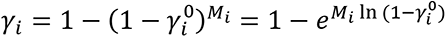 represents the probability that a susceptible agent becomes infected in region *i* per unit time. Similarly, the probability that an agent is still susceptible after *Δt* is given by lim*_δτ_*_→1_(1 − 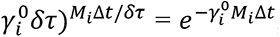, with the proportion of agents becoming infected after *Δt* being 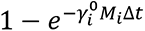.

To determine the baseline density of susceptible agents in every region, the population of susceptible agents *Ni* is incremented with:

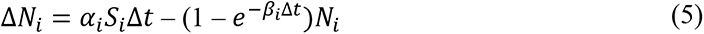

Once the system reaches its stable point (error tolerance *ε*<10^-7^), the pathogenic spread and update of *Ni* and *Mi* is given by:

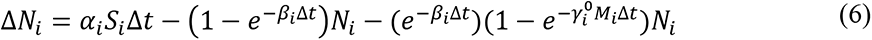

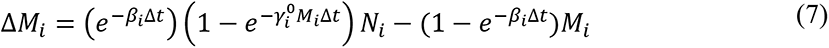

#### Propagation of normal and misfolded alpha-synuclein (module 4)

Susceptible and infected agents in region *i* either remain in region *i* or spread to other regions based on a multinomial distribution per unit time with probabilities:

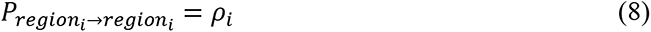

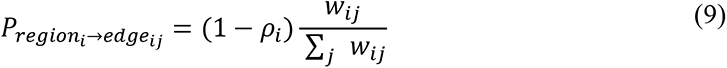

where *w_ij_* is the connection strength between regions *i* and *j* and *ρ_i_* is the probability that an agent remains in region *i*. The main analyses were performed using *ρ*=0.5 for all regions, but *ρ* values from 0.1 to 0.9 were also tested to ensure that findings did not depend solely on this parameter.

The susceptible and infected agents located inside an edge could exit the edge per unit time based on binary probabilities:

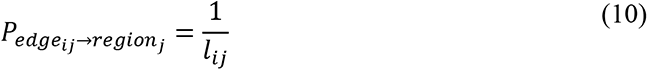

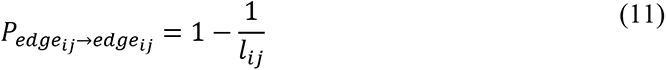

where *l_ij_* is the length of the edge between regions *i* and *j*. The increment in *N_i_* and *M_i_* in region *i* after total time *Δt* is given by:

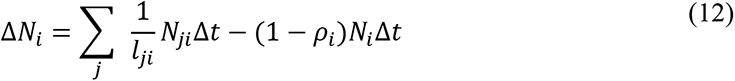

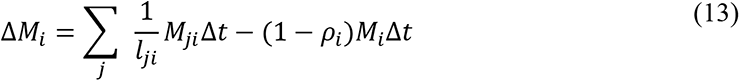

whereas the increment in the population of susceptible and infected agents inside the edge between regions *i* and *j* (*N_ij_* and *M_ij_*, respectively) after total time *Δt* is:

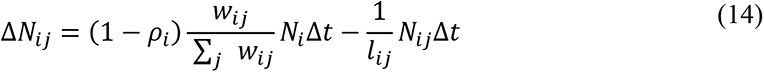

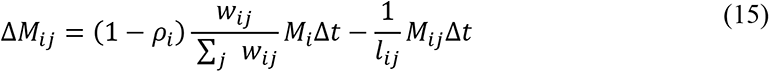

#### Emergence of simulated atrophy (module 5)

Regional atrophy was simulated as the sum of two processes: the direct toxicity resulting from the regional accumulation of infected agents and the deafferentation caused by cell death in connected regions. In region *i*, the atrophy accrual is given by:

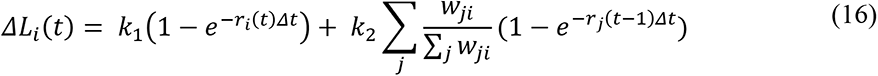

where *r_i_(t)* is the proportion of infected agents in region *i* at time *t* and 1 − *e*^−*ri*(*t*)Δ*t*^is the increment of atrophy at time *t* caused by the accumulation of alpha-synuclein pathology within *Δt*. The first term controls the direct impact of infected agents, whereas the second term weighs the increment of atrophy based on deafferentation from neighbouring regions. Each term was given a weight *k_1_* and *k_2_* of 0.5 for the main analyses, but weights varying from 0.1 to 0.9 were also tested to ensure that findings were not due to this parameter only. In other words, this module generated, at every timestep, a value of simulated atrophy for each of the 42 regions; it is this simulated atrophy that was correlated with the observed atrophy to test if the model accurately recreated the brain atrophy of iRBD.

### Statistical analysis

#### Between group differences in atrophy

For deformation-based morphometry-derived tissue deformation, two-tailed general linear models with age, sex, and site as covariates were performed to investigate the presence of significant differences between iRBD patients and controls. The Benjamini-Hochberg procedure was used to correct for the rate of false discoveries at a statistical threshold of *p*<0.05.^52^

For surface-based cortical measurements, general linear models were performed at each vertex to investigate the presence of significant differences in cortical thickness or surface area in iRBD patients compared to controls. Surface maps were smoothed using a 15-mm full-width, half-maximum kernel and age, sex, and site were entered as covariates, as well as estimated total intracranial volume for surface area. The surface correlates of motor and cognitive functioning in iRBD were also investigated in participants with available Movement Disorders Society–Unified Parkinson’s Disease Rating Scale, part III (MDS-UPDRS-III; 90 patients, 130 controls) or Montreal Cognitive Assessment (MoCA; 134 patients, 174 controls) scores. Using age, sex, and site (and total intracranial volume for cortical surface area) as covariates, vertex-based analyses were performed to investigate the vertices significantly correlated with clinical variables. Another contrast matrix was also created to identify the vertices where thickness and area correlations with the MDS-UPDRS-III or MoCA scores differed significantly between iRBD patients and controls. Statistical significance was determined by Monte Carlo simulation at a corrected threshold of *p*<0.05.

#### Replication of observed atrophy

To assess if the model recreated atrophy, the spread of pathologic alpha-synuclein was simulated in silico by injecting pathology in one region, simulating the propagation over 10,000 timesteps, and repeating the process for every region as seed. At each timestep, the model generated regional values representing the amount of simulated atrophy and the simulated number of infected and susceptible agents. To avoid interpreting any spurious overfit of the model when assessing the fit between atrophy patterns, since some regions may act as outliers due to agents being present in only a few brain regions when initiating the spread, we discarded all the timesteps where the number of infected agents in any region increased by more than 1% compared to the previous timestep.

At each timestep, Spearman’s rank correlations were used to assess the association between simulated atrophy and observed atrophy in patients. The highest correlation coefficient, if statistically significant at *p*<0.05, was considered as the peak fit. Since thickness *W*-scores correlated with region size (*r*=0.54, *p*=0.0013), the scores were divided by the region size before assessing the peak fit. For cortical surface measurements, the peak fit was also assessed over the 34 cortical regions only to ensure that the peak fit was not due to the presence of subcortical volume measurements. The *ggseg* package was used for visualization.^53^

#### Comparison with other model-derived, topological, and gene metrics

To determine that connectomics or regional vulnerability alone did not explain the atrophy patterns of iRBD, we tested whether simpler measures predicted brain atrophy as well as the complete agent-based model. These measures included (1) model-derived measurements representing the number of infected and susceptible agents at each timestep in each region; (2) network measures alone, namely node degree, node strength, node betweenness centrality and eigenvector centrality, derived from the Brain Connectivity Toolbox (www.sites.google.com/site/bctnet/);54 (3) *SNCA* and *GBA* regional expression alone. Node degree represented the number of edges (structural connections) connected to a node (region). Node strength represented the sum of the weights of the edges connected to the node. Node betweenness centrality represented the number of times a given node was found in the shortest paths linking every node pair in the network. Eigenvector centrality was a self-referential measure of centrality; nodes with high eigenvector centrality connected with other nodes that also had high eigenvector centrality. Node betweenness centrality and eigenvector centrality tested whether hub regions were more sensitive to disease.

#### Randomized null models

To test if brain connectivity and gene expression shaped the atrophy in iRBD, all fits between atrophy patterns were tested against null models in which network topology and geometry or gene expression was randomized. For the connectome, rewired and repositioned null models were used to assess network topology and/or geometry. Rewired null models are models in which structural connectivity pairs of regions were randomized while preserving the network’s original degree sequence and density. Swapping of the connectivity and distance matrices was performed using the Maslov-Sneppen algorithm in the Brain Connectivity Toolbox.^54, 55^ The randomized matrix was inserted into the model to derive a null peak fit between atrophy patterns; this process was repeated 10,000 times for generating a distribution of null peak fits, to the average of which the empirical peak fit between atrophy patterns was statistically compared. An unbiased Monte-Carlo estimate of the exact *p*-value was used to assess significance. The same steps were repeated using repositioned null models, i.e., models in which the spatial position of regions was randomized while preserving the network’s original degree sequence and connection profile. The same approach was also conducted for gene expression, where distinct null models were generated with either *SNCA* or *GBA* regional expression randomized between the 42 regions.

### Data availability

The agent-based SIR model is available at https://github.com/yingqiuz/SIR_simulator. The regional values of the tissue deformation, cortical thickness, and cortical surface area maps are available at https://github.com/srahayel/SIR-RBD.

## Results

### Demographics

Of the 443 participants, 34 did not pass deformation-based morphometry quality control, resulting in 409 participants (171 patients and 238 controls). There were no significant differences in age (iRBD: 67.7 ± 6.6 years; controls: 66.6 ± 7.9, *p*=0.11) and sex (iRBD: 83% men, controls: 77% men, *p*=0.13) between groups, but patients had higher MDS-UPDRS-III scores (*p*<0.001) and lower MoCA scores (*p*<0.001) (Table 1). Of these 409 participants, 64 did not pass FreeSurfer quality control, resulting in 345 participants (138 iRBD patients and 207 controls) for quantifying cortical thickness and surface area differences. There were no significant differences in age (iRBD: 66.2 ± 7.6 years; controls: 67.0 ± 6.3 years; *p*=0.28) and sex (iRBD: 81% men, controls: 77% men; *p*=0.34) for this sample.

**Table 1.**
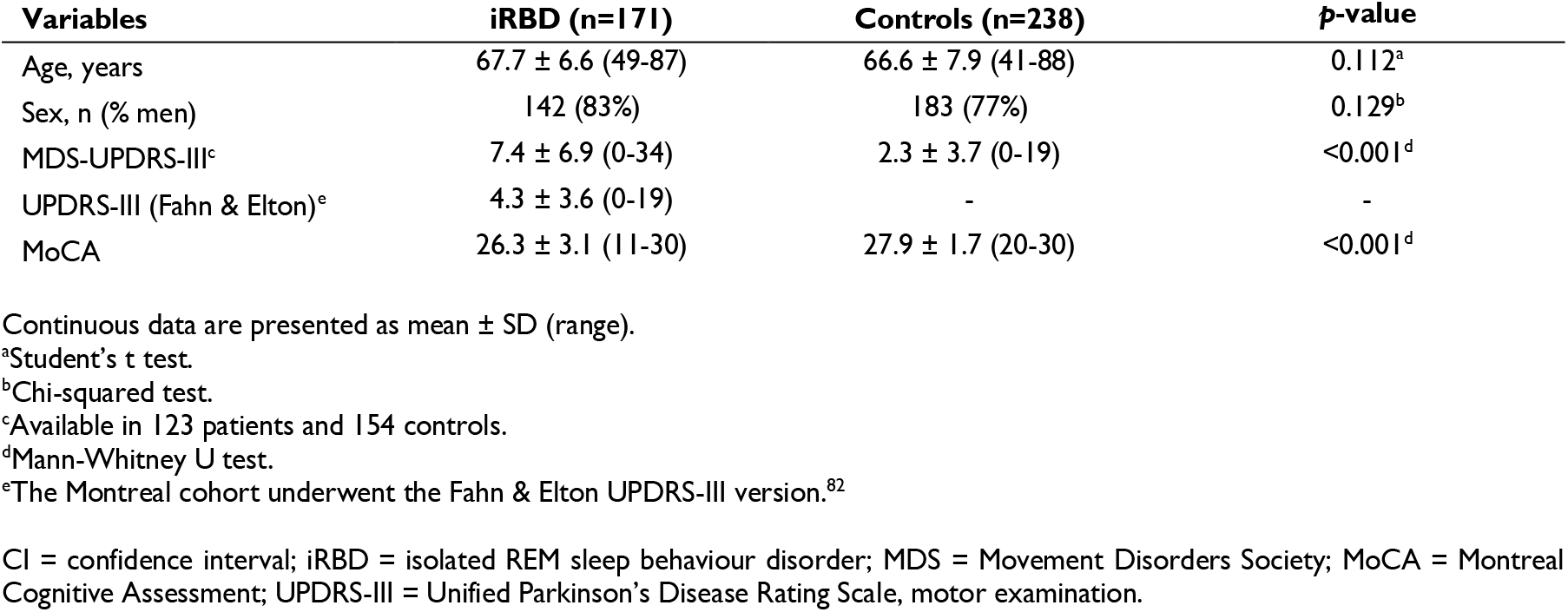
Demographics and clinical variables of iRBD patients and controls.

| Variables | iRBD (n=171) | Controls (n=238) | p-value |
| --- | --- | --- | --- |
| Age, years | 67.7 ± 6.6 (49-87) | 66.6 ± 7.9 (41-88) | 0.112<sup>a</sup> |
| Sex, n (% men) | 142 (83%) | 183 (77%) | 0.129<sup>b</sup> |
| MDS-UPDRS-III<sup>c</sup> | 7.4 ± 6.9 (0-34) | 2.3 ± 3.7 (0-19) | <0.001<sup>d</sup> |
| UPDRS-III (Fahn & Elton)<sup>e</sup> | 4.3 ± 3.6 (0-19) | - | - |
| MoCA | 26.3 ± 3.1 (11-30) | 27.9 ± 1.7 (20-30) | <0.001<sup>d</sup> |

Continuous data are presented as mean ± SD (range).
<sup>a</sup>Student's t test.
<sup>b</sup>Chi-squared test.
<sup>c</sup>Available in 123 patients and 154 controls.
<sup>d</sup>Mann-Whitney U test.
<sup>e</sup>The Montreal cohort underwent the Fahn & Elton UPDRS-III version.<sup>82</sup>
CI = confidence interval; iRBD = isolated REM sleep behaviour disorder; MDS = Movement Disorders Society; MoCA = Montreal Cognitive Assessment; UPDRS-III = Unified Parkinson's Disease Rating Scale, motor examination.

### iRBD patients show brain atrophy

We investigated if this iRBD cohort showed brain atrophy compared to controls. In terms of deformation-based morphometry-derived tissue deformation, patients had decreased volume in the left middle temporal cortex, cuneus, lingual gyrus, fusiform gyrus, banks of the superior temporal sulcus and the pericalcarine area, and increased volume in the insula compared to controls; in the right hemisphere, decreased volume was found in the precentral, supramarginal, superior and middle temporal, lingual, and cuneus regions. However, only the left middle temporal region was significant after correction (*p_FDR_*=0.045) (Fig. 2A and Supplementary Table 2).

**Figure 2.**
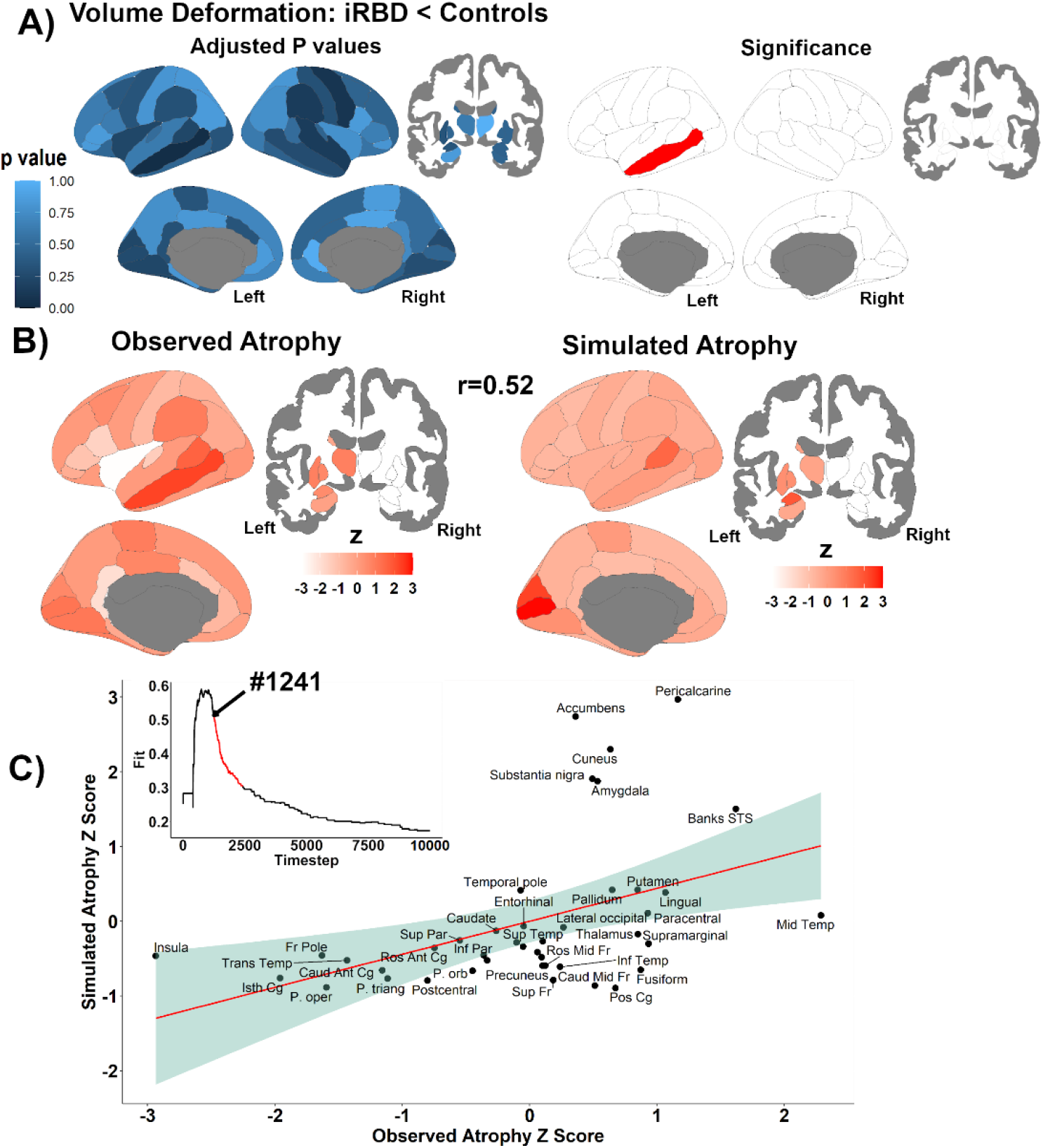
The model recreates the pattern of tissue deformation of iRBD. **(A)** Patients with iRBD showed significant volume decreases compared to controls (left: spatial distribution of FDR-adjusted *p*-values, right: significant differences after FDR-correction). **(B)** The pattern of volume loss in iRBD (left) was recreated by the model (right). For visualization purposes, atrophy *W*-scores were *z*-scored to ease comparability of scales; positive *z* scores represented greater atrophy. **(C)** The subplot shows the progression of the fit between atrophy patterns until the peak at timestep #1241 (arrow). The main scatterplot shows the *z*-scored values of observed and simulated atrophy at the peak fit for the 42 regions. Scales were adjusted such that higher scores represented greater atrophy. FDR = false discovery rate; iRBD = isolated REM sleep behaviour disorder.

For cortical thickness, patients showed significant thinning compared to controls in two clusters in the left hemisphere, namely one posterior cluster that included the posterior temporal and inferior parietal cortices and another cluster that extended from the dorsolateral prefrontal cortex to the orbitofrontal cortex, and in one cluster in the right hemisphere, which included the posterior temporal and lateral occipital cortices (Fig. 3A and Table 2). Compared to controls, patients also had significantly increased cortical surface area in the left inferior temporal cortex and sulcus that extended to the entorhinal cortex (Fig. 3B and Table 2). These findings demonstrate the presence of brain atrophy in iRBD.

**Figure 3.**
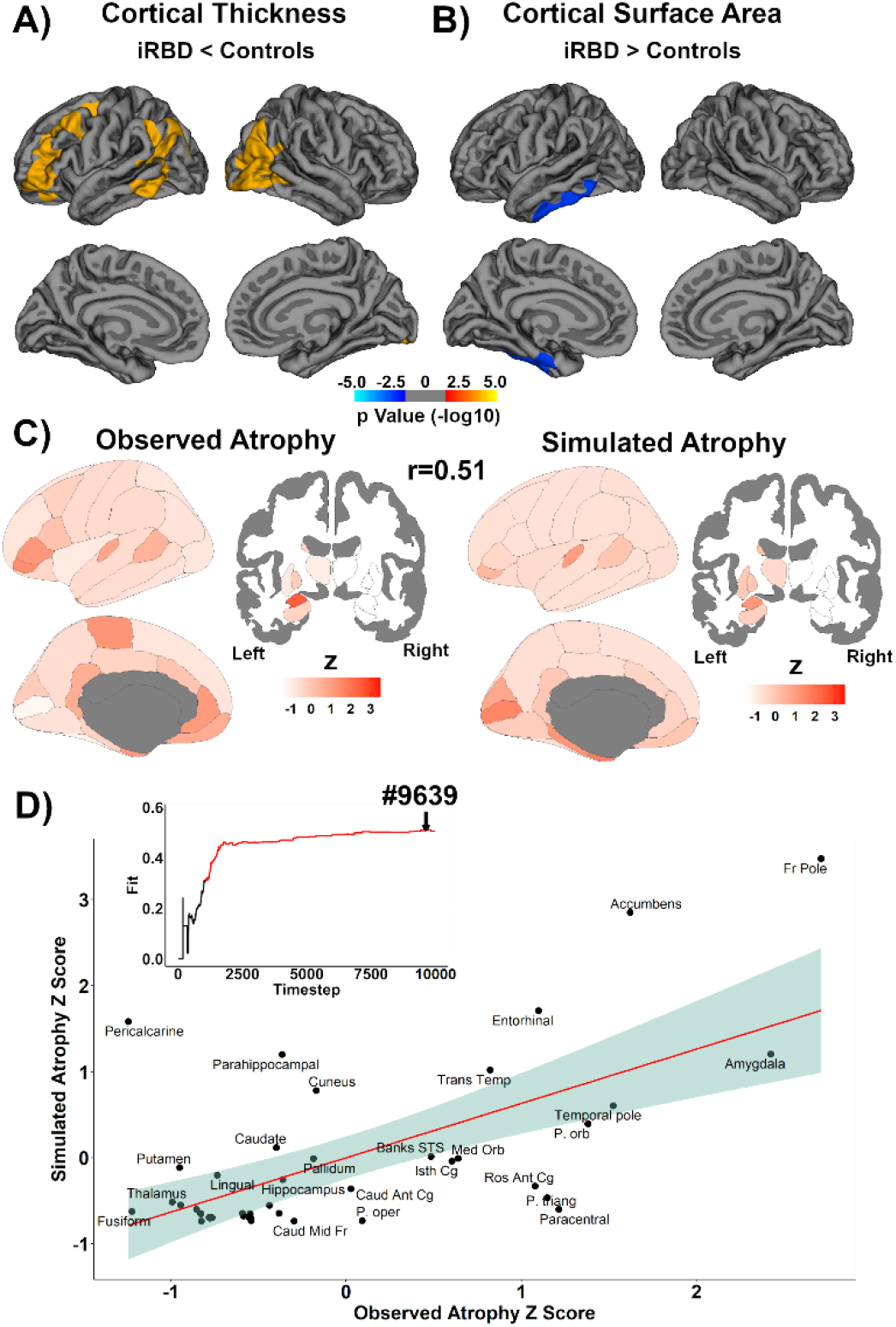
The model recreates the pattern of cortical thinning of iRBD. Patients with iRBD showed cortical thinning **(A)** and increased surface area **(B)** compared to controls. The colour bar indicates the statistical significance on a logarithmic scale of *p*- values (-log10), with red-yellow areas showing significant decreases in iRBD and blue areas showing increases in iRBD. **(C)** The pattern of cortical thinning in iRBD (left) was recreated by the model (right). For visualization purposes, atrophy *W*-scores were *z*-scored to ease comparability of scales; positive *z* scores represented greater atrophy; positive *z* scores represent greater atrophy. **(D)** The subplot shows the progression of the fit between atrophy patterns until the peak at timestep 9,639 (arrow). The main scatterplot shows the *z*-scored values of observed and simulated atrophy at the peak fit for the 41 regions. Scales were adjusted such that higher scores represented greater atrophy. iRBD = isolated REM sleep behaviour disorder.

**Table 2.**
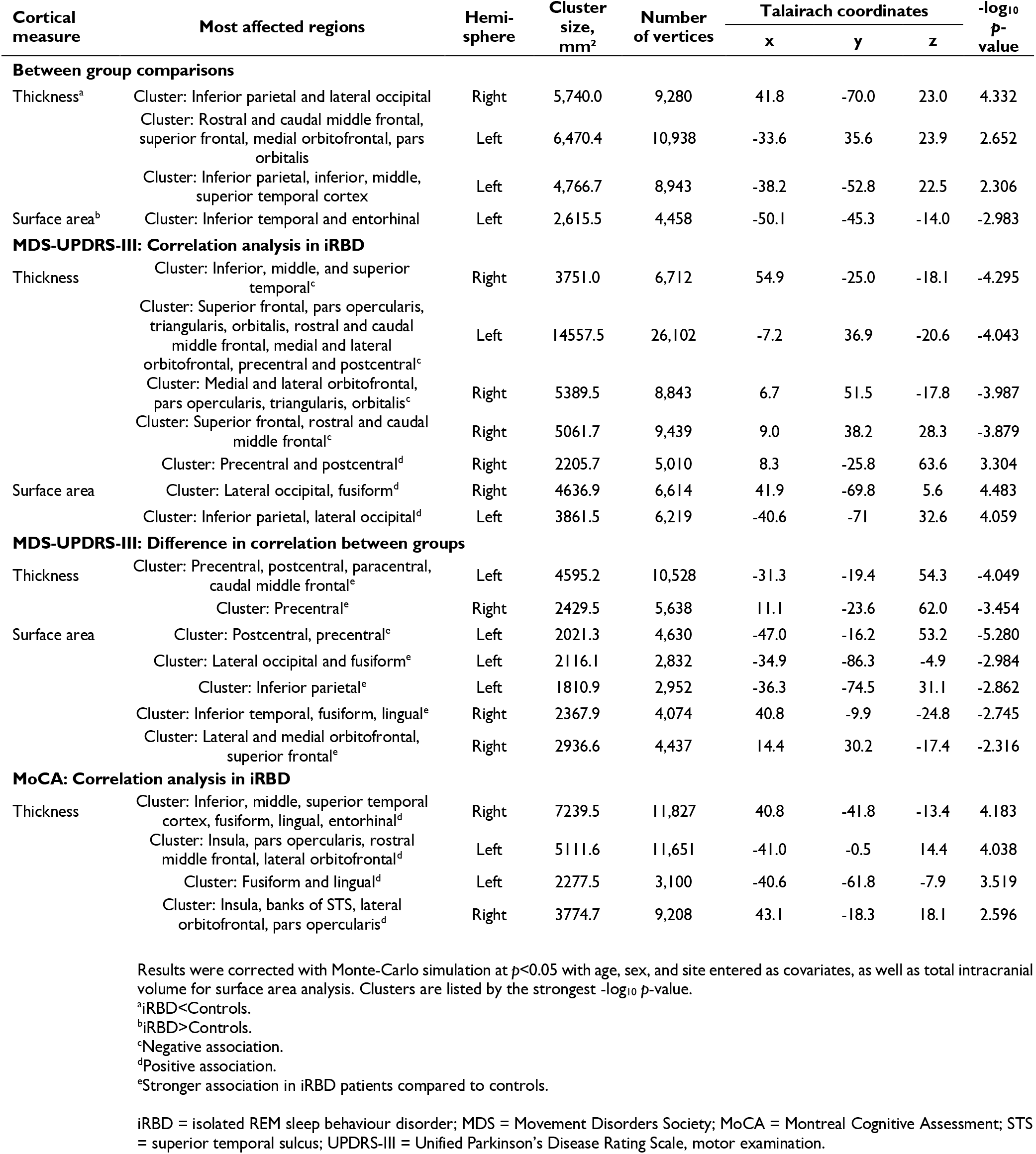
Results of vertex-based cortical analyses between iRBD patients and controls.

### Brain atrophy in iRBD is associated with motor and cognitive functioning

In iRBD patients, higher MDS-UPDRS-III scores were associated with cortical thinning in the bilateral frontal cortex and the right temporal cortex, and with increased thickness in the right sensorimotor cortex (Fig. 4B and Table 2). Higher MDS-UPDRS-III scores were also associated with increased cortical surface area in the bilateral occipital cortex, the left inferior parietal cortex, and the right posterior temporal cortex (Fig. 4B and Table 2). The associations with MDS-UPDRS-III scores were significantly different between iRBD patients and controls, with the correlation being stronger in patients in the bilateral sensorimotor cortex for thickness and in the frontopolar, sensorimotor, occipital, inferior parietal, and lingual and fusiform cortices for surface area (Fig. 4B and Table 2). For the MoCA, lower scores were associated with cortical thinning in the bilateral insula, the right temporal cortex, and the left posterior temporal cortex (Fig. 4C and Table 2). MoCA scores were not associated with cortical surface area in iRBD patients and there were no differences in correlation slopes between patients and controls.

**Figure 4.**
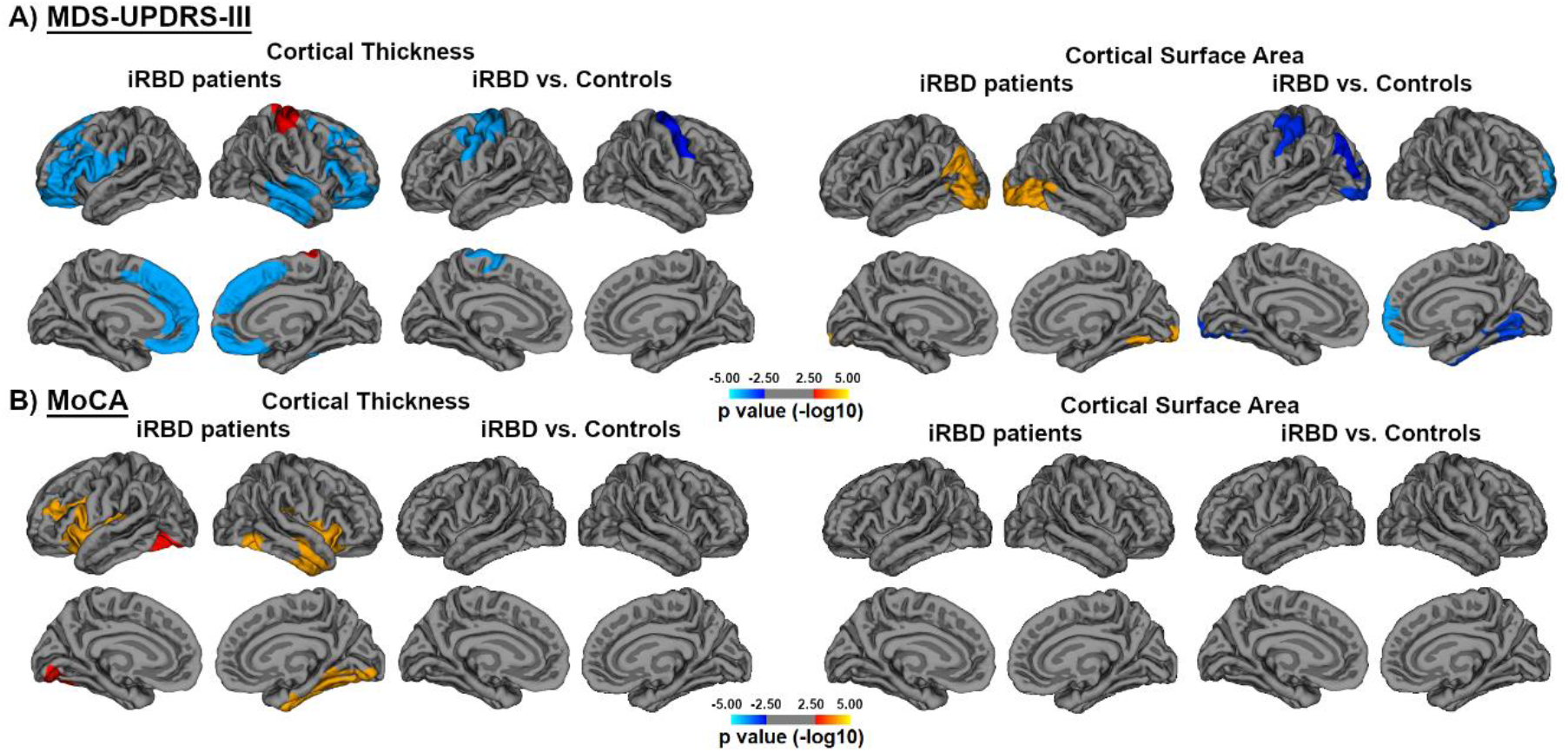
Correlation analyses between cortical surface and motor and cognitive variables. Results of the general linear models showing the vertices where a significant correlation was found with the MDS-UPDRS-III **(A)** and MoCA **(B)** in iRBD patients and the vertices where the slopes between the structural metric and these scores significantly differed between patients and controls. The colour bar indicates the statistical significance on a logarithmic scale of *p*-values (-log10; ±1.3 corresponding to *p*<0.05 corrected for multiple comparisons by Monte Carlo cluster-wise simulation), with red-yellow areas showing positive associations in iRBD and blue areas showing negative associations in iRBD. For comparisons of correlation, blue clusters represent stronger correlations in iRBD compared to controls. iRBD = isolated REM sleep behaviour disorder; MDS = Movement Disorders Society; MoCA = Montreal Cognitive Assessment; UPDRS-III = Unified Parkinson’s Disease Rating Scale, part III.

### The SIR model recreates the atrophy of iRBD

The agent-based model was then applied to simulate alpha-synuclein spread and generate patterns of simulated atrophy in every region. We found that the model recreated the deformation-based morphometry-derived tissue deformation pattern, with the peak fit reaching *r*=0.52 (*p*<0.0005) when seeding from the banks of superior temporal sulcus at a connection density of 40% (Fig. 2B). The fit between atrophy patterns increased gradually with each timestep to reach a peak, followed by a decline (Fig. 2C). At the peak, the simulated atrophy was most prominent in the pericalcarine cortex, accumbens, cuneus, substantia nigra, amygdala, and the banks of the superior temporal sulcus (Fig. 2C). At later timesteps, the correlation declined but simulated atrophy was seen in putamen and lingual gyrus. The model also recreated atrophy at the lower connection densities (*r*=0.44 at 35%, *r*=0.50 at 30%, *r*=0.36 at 25%) (Supplementary Fig. 1).

Similarly, the model recreated cortical atrophy, derived from FreeSurfer, with peak fits reaching *r*=0.51 (*p*=0.0007) for cortical thickness (Fig. 3C) and *r*=0.43 (*p*=0.006) for cortical surface area. However, when assessing the fit over the 34 cortical regions only (without the subcortical measurements), only thickness remained significant (*r_thickness_*=0.48, *p*=0.004; *r_area_*=0.28, *p*=0.11). The fit for cortical thickness increased gradually and reached its peak once the system had attained the equilibrium state; at this timestep, the simulated atrophy was primarily found in the frontal pole, amygdala, accumbens, and entorhinal cortex (Fig. 3D). The fit was also significant at lower connection densities (*r*=0.50 at 35%, *r*=0.49 at 30%, *r*=0.48 at 25%).

To confirm that our findings were not caused by site-specific variability, the tissue deformation and cortical thickness *W*-scores were harmonized using ComBat^44, 45^; the model recreated the patterns just described in a similar way (*r*=0.515, *p*=5.69x10^-4^ for tissue deformation, *r*=0.523, *p*=5.50x10^-4^ for cortical thickness). In addition, the model also recreated atrophy when using finer parcellations of 65 and 119 regions (Supplementary Fig. 2), with various spreading rates of agents (Supplementary Fig. 3), and with different weights given to misfolded alpha-synuclein accumulation versus deafferentation in the simulated atrophy ratio, with higher peak fits obtained when deafferentation was given the larger weight (Supplementary Fig. 4). Taken together, these results demonstrate that volume and cortical thickness atrophy in iRBD can be recreated based on agent-based modelling utilizing connectivity and gene expression for generation of the model.

### Simulated atrophy outperforms gene and network metrics

To ascertain whether brain connectivity or gene expression alone could recreate the atrophy pattern as well as the full model, we also tested several network-based and other model-derived measures. For every connection density, we found that the simulated atrophy from the agent-based model measure always yielded the highest peak fits (*r*=0.52, *p*<0.0005 at 40%), followed by the number of susceptible agents (*r*=0.32, *p*=0.039) (Fig. 5). None of the other measures, namely the number of infected agents or those describing the network’s topology, associated significantly with measured atrophy (Fig. 5). As for gene expression, the deformation-based morphometry-derived tissue deformation was associated with neither *SNCA* (*r*=0.24, *p*=0.12) nor *GBA* (*r*=0.18, *p*=0.24) regional expression. This demonstrates that brain connectivity or gene expression alone cannot predict atrophy in iRBD, and that the full agent-based SIR model taking into account gene expression, connectivity, and deafferentation provides the best fit to the measured deformation.

**Figure 5.**
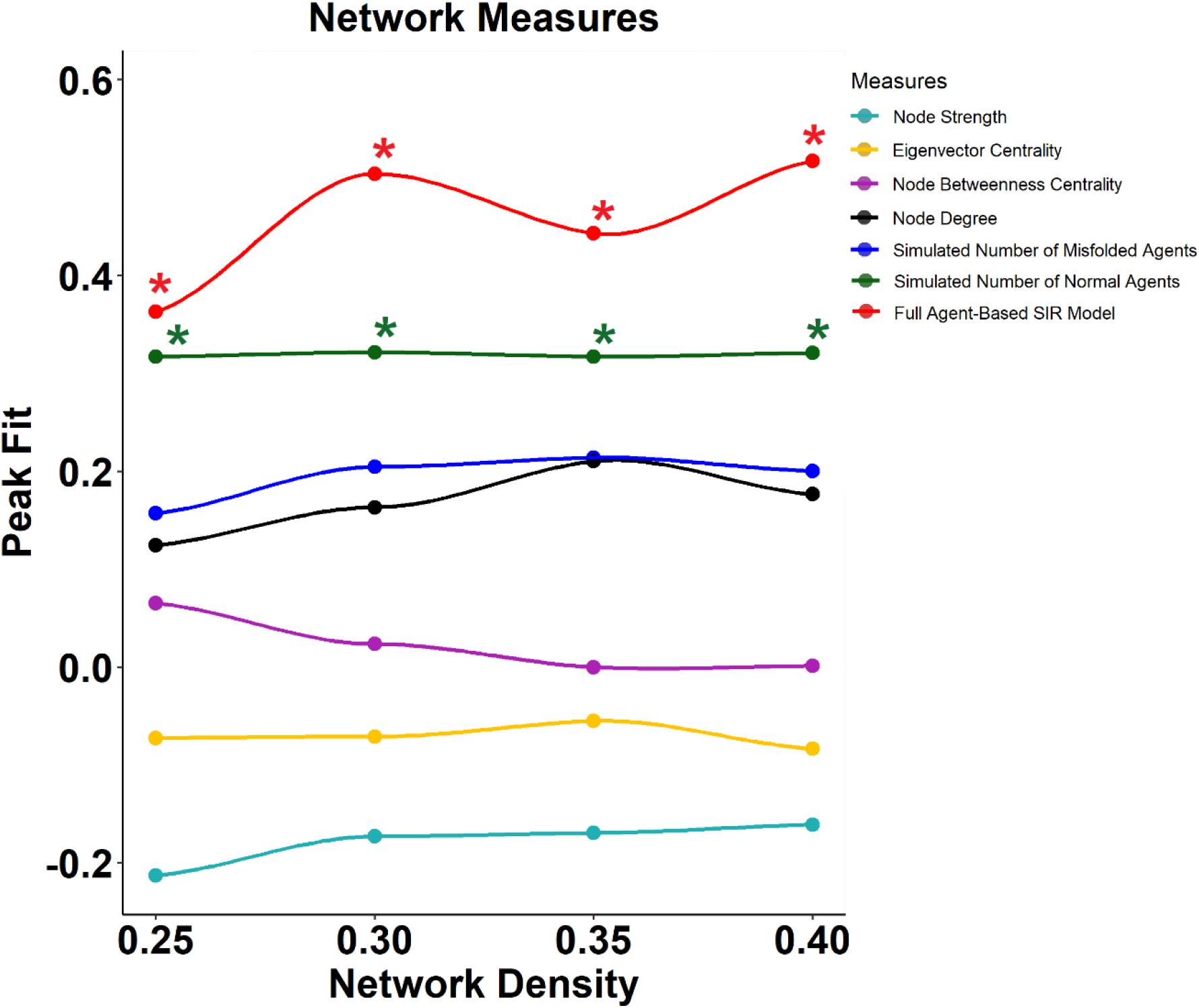
Simulated atrophy outperforms network and other model-derived measures. The highest peak fits found when recreating the deformation-based morphometry-derived tissue deformation pattern were found when using the simulated atrophy measure generated by the full agent-based SIR model. Asterisks represent the significant correlations between the observed pattern of atrophy and the different network and model-derived measures. SIR = Susceptible-Infected-Removed.

### Connectivity and gene expression shape alpha-synuclein spread

We further investigated the importance of connectome architecture and gene expression by generating several null models. The peak fits of the SIR model were compared to the peak fits observed in null distributions derived from simulations where either gene expression or network architecture were randomly shuffled across regions. We observed that randomizing *SNCA* and *GBA* expression levels (i.e., alpha-synuclein synthesis and clearance) significantly disrupted the model fit at a 40% connection density; however, randomizing the *SNCA* expression did not disrupt the fit at lower connection densities (Fig. 6). The randomization of connectivity was performed using two different types of null models to assess the impact of network topology and/or geometry on alpha-synuclein spread. In both cases, randomizing the connectome’s architecture disrupted the model fit (Fig. 6), indicating that both the brain’s structural connectivity pattern and the physical constraints imposed on the connectome contribute to shaping atrophy. Taken together, this demonstrates that connectivity and gene expression combine to shape brain atrophy of iRBD.

**Figure 6.**
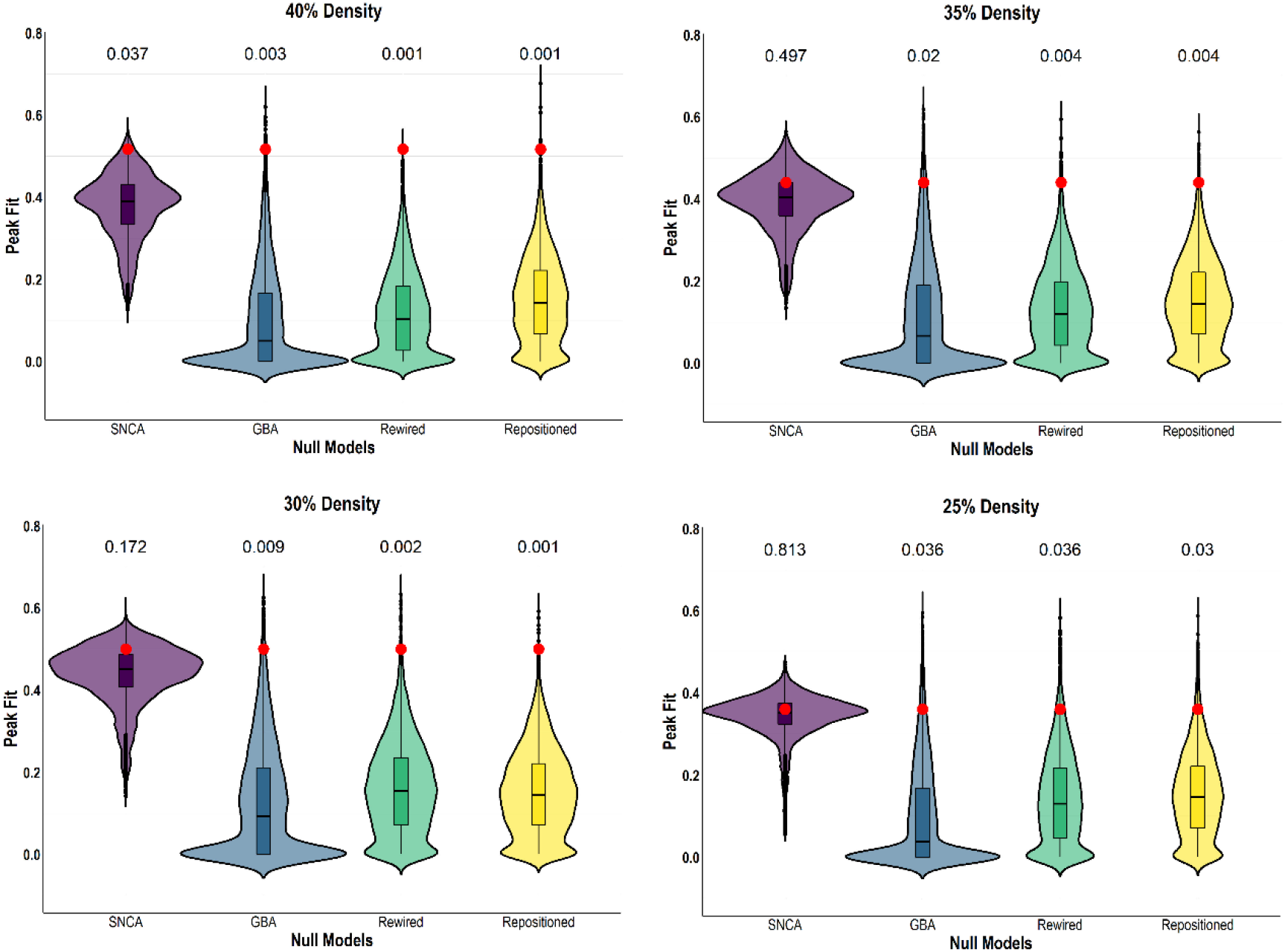
Connectivity and gene expression shape the deformation-based morphometry-derived atrophy pattern. Violin plots comparing the original peak fits between atrophy patterns (red dots) to the average peak fits derived from null models in which network topology (“Rewired”), network geometry (“Repositioned”), *SNCA* expression (“SNCA”), and *GBA* expression (“GBA”) was randomly shuffled across regions. Unbiased Monte-Carlo estimates of the exact *p*-value were computed and reported above the plots. Comparisons with null models were performed for each connection density.

## Discussion

Isolated RBD is associated with brain volume and cortical thickness atrophy.^6–8, 56–61^ Patients also present with high rates of positivity to pathologic alpha-synuclein in tissue biopsies.^62–65^ However, a mechanistic understanding of how alpha-synuclein pathology may relate to the patterns of brain atrophy in iRBD remains unknown. Here, we quantified atrophy in the largest multicentric cohort of polysomnography-confirmed iRBD patients with T1-weighted MRI acquired to date and applied the agent-based SIR model to test whether prion-like and regional vulnerability factors recreated the atrophy in iRBD. We demonstrated that the computational simulation of atrophy based on the connectome’s architecture and gene expression of *SNCA* and *GBA* did recreate the atrophy seen in iRBD patients. Since most of the atrophy in iRBD associates with cognitive impairment,^7^ and given that atrophy predicts dementia with Lewy bodies in iRBD,^8^ this study sheds light on the specific mechanisms underlying the development of dementia with Lewy bodies in this population.

The agent-based SIR model simulates the spread of alpha-synuclein based on brain connectivity and the expression of genes involved in alpha-synuclein synthesis and clearance.^21^ This model recreates the atrophy seen in Parkinson’s disease^21^ and frontotemporal dementia,^47^ and the spatiotemporal patterns of pathologic alpha-synuclein quantified in non-transgenic mice injected with preformed fibrils into either the striatum, nucleus accumbens or hippocampus.^22^ In the current work, we show that the atrophy in iRBD also follows the constraints imposed by the connectome’s architecture and the gene expression of *SNCA* and *GBA*. The impact of these factors was decisive, as demonstrated by the model’s inability to recreate atrophy if either connectivity or gene expression were randomized. The influence of connectivity is in line with several studies showing that cerebral connectivity forecasts the atrophy seen in neurodegenerative diseases,^66–68^ including Parkinson’s disease.^20, 21^ The influence of *SNCA* and *GBA* gene expression also agrees with mutations in these genes being significant risk factors for Parkinson’s disease and dementia with Lewy bodies.^69–71^ We observed that, whereas randomizing *GBA* expression always interfered with the model’s ability to recreate atrophy, the randomization of *SNCA* expression only led to a disruption at the 40% connection density, suggesting that *GBA* expression exerts an effect on the spread (and hence atrophy) that is more consistent than *SNCA*. The reason for this difference is unclear but does not seem related to different gene expression levels used by the model (*SNCA*: 0.17 ± 0.66 (range: 2.91) versus *GBA*: 0.03 ± 0.51 (range: 2.95), *p*=0.28). Another possibility is that the 90 additional connections appearing in the 40% density connectome were particularly influential for *SNCA* to exert its effect on atrophy. Further studies should therefore investigate more closely the effect of gene expression on atrophy in iRBD. Nonetheless, this is consistent with our previous work showing that atrophy in Parkinson’s disease is shaped by local concentration of alpha-synuclein and connectivity, supporting the prion model.^21^

However, although the model recreated atrophy, the visual inspection of observed and simulated atrophy measurements at the peak fit showed some inconsistencies. For instance, while the middle temporal gyrus was the region showing the greatest tissue deformation in iRBD, the amount of simulated atrophy was modest. This could suggest that proteins other than alpha-synuclein, disease-related changes in connectivity, regional vulnerability to alpha-synuclein accumulation unaccounted for in the model, and/or other mechanistic explanations may be involved in the observed changes in the middle temporal gyrus. However, in contrast, neighbouring regions such as the amygdala and the entorhinal cortex, which were reported to show high burden of Lewy pathology in brains of Lewy body patients,^72^ were also among the regions showing the highest amount of simulated atrophy in our patterns.

Another novelty of this study is the comprehensive assessment of brain morphology in a large multicentric cohort of iRBD patients with polysomnography and T1-weighted MRI acquisition. We found that iRBD patients had volume atrophy in the middle temporal gyrus and cortical thinning in the frontal, posterior temporal, occipital, and inferior parietal cortices, which is in line with earlier findings in smaller iRBD cohorts.^28^ Morphological changes related to clinical changes, with the severity of parkinsonism being associated with extensive cortical thinning, thickening of the sensorimotor cortex, and increased area in posterior regions. Whereas the paracentral, sensorimotor, and superior parietal areas have been associated with motor deficits in iRBD,^6, 59^ other regions such as the frontal and temporal cortices were not expected to be involved based on previous work. However, this frontotemporal pattern has already been documented in Parkinson’s disease^73^ and our findings here may represent premorbid neurodegenerative changes typical of overt Parkinson’s disease. As for the MoCA correlates, the pattern was in line with what was previously observed for global cognitive performance in Parkinson’s disease^73^ and included the anterior insula, which was shown to be particularly vulnerable in synucleinopathies affecting cognition.^74–77^

This study has some limitations. First, the brainstem nuclei involved in REM sleep motor atonia^78^ were not included in our analyses. This is due to the difficulty in imaging both brain atrophy and connectivity in these structures. Second, scans were acquired using different acquisition parameters at different sites. In the current work, site effects were regressed out from atrophy measurements during *W*-scoring and entered as covariate in neuroimaging analyses. The use of harmonized *W*-scores using a batch-correcting tool validated for neuroimaging data yielded the same results. Third, alpha-synuclein spread was simulated on a healthy structural connectome and transcriptome. White matter abnormalities and topological disorganization of grey matter have been reported in iRBD,^8, 58, 79, 80^ as well as a genetic makeup more complex than the sole effects of *SNCA* and *GBA*.^81^ Once these changes are more thoroughly understood in iRBD, they can be implemented in the model.

In summary, atrophy in iRBD patients can be recreated using a combination of agent-based modelling, structural connectomics, and gene expression. The agent-based SIR model may provide a way to test new research hypotheses for the purpose of slowing or stopping the spread of pathologic alpha-synuclein in the brain.

## Acknowledgements

Shady Rahayel reports a scholarship from the Fonds de recherche du Québec – Santé. Jean-François Gagnon holds a Canada Research Chair in Cognitive Decline in Pathological Aging. Kaylena Ehgoetz Martens reports a scholarship from Parkinson Canada and Sydney Fellowship. Data used in this article included data from the Parkinson’s Progression Markers Initiative database (https://www.ppmi-info.org), a public-private partnership, funded by the Michael J. Fox Foundation for Parkinson’s Research.

## Appendix 1

List of the contributors involved in the ICEBERG Study Group:

Steering committee: Marie Vidailhet, MD, PhD, (Pitié-Salpêtrière Hospital, Paris, principal investigator of ICEBERG), Jean-Christophe Corvol, MD, PhD (Pitié-Salpêtrière Hospital, Paris, scientific lead), Isabelle Arnulf, MD, PhD (Pitié-Salpêtrière Hospital, Paris, member of the steering committee), Stéphane Lehericy, MD, PhD (Pitié-Salpêtrière Hospital, Paris, member of the steering committee);

Clinical data: Marie Vidailhet, MD, PhD, (Pitié-Salpêtrière Hospital, Paris, coordination), Graziella Mangone, MD, PhD (Pitié-Salpêtrière Hospital, Paris, co-coordination), Jean-Christophe Corvol, MD, PhD (Pitié-Salpêtrière Hospital, Paris), Isabelle Arnulf, MD, PhD (Pitié-Salpêtrière Hospital, Paris), Sara Sambin, MD (Pitié-Salpêtrière Hospital, Paris), Jonas Ihle, MD (Pitié-Salpêtrière Hospital, Paris), Caroline Weill, MD, (Pitié-Salpêtrière Hospital, Paris), David Grabli, MD, PhD (Pitié-Salpêtrière Hospital, Paris); Florence Cormier-Dequaire, MD (Pitié-Salpêtrière Hospital, Paris); Louise Laure Mariani, MD, PhD (Pitié-Salpêtrière Hospital, Paris), Bertrand Degos, MD, PhD (Avicenne Hospital, Bobigny);

Neuropsychological data: Richard Levy, MD (Pitié-Salpêtrière Hospital, Paris, coordination), Fanny Pineau, MS (Pitié-Salpêtrière Hospital, Paris, neuropsychologist), Julie Socha, MS (Pitié-Salpêtrière Hospital, Paris, neuropsychologist), Eve Benchetrit, MS (La Timone Hospital, Marseille, neuropsychologist), Virginie Czernecki, MS (Pitié-Salpêtrière Hospital, Paris, neuropsychologist), Marie-Alexandrine, MS (Pitié-Salpêtrière Hospital, Paris, neuropsychologist);

Eye movement: Sophie Rivaud-Pechoux, PhD (ICM, Paris, coordination); Elodie Hainque, MD, PhD (Pitié-Salpêtrière Hospital, Paris);

Sleep assessment: Isabelle Arnulf, MD, PhD (Pitié-Salpêtrière Hospital, Paris, coordination), Smaranda Leu Semenescu, MD (Pitié-Salpêtrière Hospital, Paris), Pauline Dodet, MD (Pitié-Salpêtrière Hospital, Paris);

Genetic data: Jean-Christophe Corvol, MD, PhD (Pitié-Salpêtrière Hospital, Paris, coordination), Graziella Mangone, MD, PhD (Pitié-Salpêtrière Hospital, Paris, co-coordination), Samir Bekadar, MS (Pitié-Salpêtrière Hospital, Paris, biostatistician), Alexis Brice, MD (ICM, Pitié-Salpêtrière Hospital, Paris), Suzanne Lesage, PhD (INSERM, ICM, Paris, genetic analyses);

Metabolomics: Fanny Mochel, MD, PhD (Pitié-Salpêtrière Hospital, Paris, coordination), Farid Ichou, PhD (ICAN, Pitié-Salpêtrière Hospital, Paris), Vincent Perlbarg, PhD, Pierre and Marie Curie University), Benoit Colsch, PhD (CEA, Saclay), Arthur Tenenhaus, PhD (Supelec, Gif-sur-Yvette, data integration);

Brain MRI data: Stéphane Lehericy, MD, PhD (Pitié-Salpêtrière Hospital, Paris, coordination), Rahul Gaurav, MS, (Pitié-Salpêtrière Hospital, Paris, data analysis), Nadya Pyatigorskaya, MD, PhD, (Pitié-Salpêtrière Hospital, Paris, data analysis); Lydia Yahia-Cherif, PhD (ICM, Paris, Biostatistics), Romain Valabregue, PhD (ICM, Paris, data analysis), Cécile Galléa, PhD (ICM, Paris);

Datscan imaging data: Marie-Odile Habert, MCU-PH (Pitié-Salpêtrière Hospital, Paris, coordination);

Voice recording: Dijana Petrovska, PhD (Telecom Sud Paris, Evry, coordination), Laetitia Jeancolas, MS (Telecom Sud Paris, Evry);

Study management: Vanessa Brochard (Pitié-Salpêtrière Hospital, Paris, coordination), Alizé Chalançon (Pitié-Salpêtrière Hospital, Paris, Project manager), Carole Dongmo-Kenfack (Pitié-Salpêtrière Hospital, Paris, clinical research assistant); Christelle Laganot (Pitié-Salpêtrière Hospital, Paris, clinical research assistant), Valentine Maheo (Pitié-Salpêtrière Hospital, Paris, clinical research assistant).

## Supplementary material

**Supplementary Table 1.** Demographics of the different cohorts.

| Cohort | Group | Total number | Age (SD) | Sex, n (% men) | MDS-UPDRS-III | MoCA |
| --- | --- | --- | --- | --- | --- | --- |
| Paris | iRBD | 59 | 67.42 ± 6.16 | 52 (88) | 8.47 ± 6.68 | 26.78 ± 2.79 |
| Paris | Controls | 57 | 65.94 ± 8.51 | 45 (79) | 3.98 ± 4.77 | 27.51 ± 2.16 |
| Montreal | iRBD | 48 | 65.83 ± 6.43 | 37 (77) | 4.31 ± 3.62<sup>a</sup> | 25.93 ± 2.72 |
| Montreal | Controls | 35 | 64.97 ± 7.13 | 21 (60) | - | 28.17 ± 1.40 |
| Sydney | iRBD | 30 | 68.42 ± 7.24 | 27 (90) | 11.58 ± 7.45 | 26.42 ± 3.34 |
| Sydney | Controls | 26 | 70.81 ± 6.22 | 17 (65) | - | - |
| Aarhus | iRBD | 18 | 68.72 ± 8.22 | 15 (83) | 1.11 ± 1.45 | 27.11 ± 1.97 |
| Aarhus | Controls | 20 | 67.90 ± 5.76 | 15 (75) | - | 27.05 ± 2.12 |
| PPMI cohort | iRBD | 27 | 70.86 ± 5.32 | 21 (78) | 5.11 ± 5.64 | 25.18 ± 4.51 |
| PPMI cohort | Controls | 123 | 66.07 ± 8.18 | 97 (79) | 1.43 ± 2.46 | 28.15 ± 1.14 |

Data are presented as mean ± SD.
<sup>a</sup>Fahn & Elton UPDRS-III version.<sup>82</sup>
iRBD = isolated REM sleep behaviour disorder; MDS = Movement Disorders Society; MoCA = Montreal Cognitive Assessment; PPMI = Parkinson's Progression Markers Initiative; SD = standard deviation; UPDRS-III = Unified Parkinson's Disease Rating Scale, motor examination.

**Supplementary Table 2.** Results of deformation-based morphometry between iRBD patients and controls.

| Region | iRBD (mean ± SD) | Controls (mean ± SD) | Beta coefficient | p-value | Adjusted p-value for FDR |
| --- | --- | --- | --- | --- | --- |
| **Left hemisphere** |  |  |  |  |  |
| Middle temporal cortex<sup>a</sup> | 1.008 (0.053) | 1.019 (0.050) | -0.016 | 0.001 | 0.045 |
| Cuneus<sup>b</sup> | 1.007 (0.079) | 1.014 (0.082) | -0.018 | 0.030 | 0.274 |
| Lingual gyrus<sup>b</sup> | 1.015 (0.063) | 1.022 (0.066) | -0.014 | 0.033 | 0.274 |
| Fusiform gyrus<sup>b</sup> | 1.024 (0.085) | 1.028 (0.079) | -0.014 | 0.032 | 0.274 |
| Banks STS<sup>b</sup> | 0.976 (0.081) | 0.988 (0.073) | -0.017 | 0.026 | 0.274 |
| Insula<sup>b</sup> | 1.013 (0.053) | 0.999 (0.044) | 0.011 | 0.030 | 0.274 |
| Pericalcarine<sup>b</sup> | 0.960 (0.082) | 0.973 (0.090) | -0.018 | 0.043 | 0.286 |
| Supramarginal | 1.004 (0.052) | 1.010 (0.050) | -0.010 | 0.062 | 0.325 |
| Isthmus cingulate | 0.991 (0.055) | 0.979 (0.057) | 0.011 | 0.066 | 0.326 |
| Frontal pole | 1.046 (0.052) | 1.033 (0.056) | 0.009 | 0.112 | 0.428 |
| Precentral | 1.029 (0.046) | 1.031 (0.044) | -0.007 | 0.110 | 0.428 |
| Paracentral | 1.008 (0.081) | 1.017 (0.079) | -0.013 | 0.119 | 0.428 |
| Caudal anterior cingulate | 0.979 (0.063) | 0.970 (0.060) | 0.010 | 0.127 | 0.428 |
| Lateral occipital | 1.029 (0.039) | 1.032 (0.047) | -0.007 | 0.124 | 0.428 |
| Putamen | 0.916 (0.062) | 0.925 (0.058) | -0.009 | 0.099 | 0.428 |
| Amygdala | 0.993 (0.082) | 0.996 (0.071) | -0.011 | 0.106 | 0.428 |
| Caudal middle frontal | 1.019 (0.058) | 1.025 (0.064) | -0.009 | 0.148 | 0.435 |
| Entorhinal | 1.047 (0.121) | 1.044 (0.106) | -0.012 | 0.208 | 0.452 |
| Caudate | 1.027 (0.120) | 1.013 (0.109) | 0.015 | 0.204 | 0.452 |
| Pallidum | 0.913 (0.064) | 0.923 (0.061) | -0.007 | 0.221 | 0.453 |
| Inferior temporal | 1.047 (0.090) | 1.043 (0.078) | -0.007 | 0.230 | 0.454 |
| Lateral orbitofrontal | 1.001 (0.072) | 0.999 (0.071) | -0.006 | 0.252 | 0.480 |
| Temporal pole | 1.015 (0.089) | 1.014 (0.077) | -0.008 | 0.262 | 0.488 |
| Transverse temporal | 1.004 (0.083) | 0.990 (0.081) | 0.009 | 0.292 | 0.515 |
| Thalamus | 0.959 (0.059) | 0.960 (0.055) | -0.005 | 0.384 | 0.614 |
| Accumbens area | 1.012 (0.070) | 1.008 (0.064) | -0.005 | 0.387 | 0.614 |
| Superior temporal | 1.005 (0.041) | 1.005 (0.045) | -0.004 | 0.399 | 0.621 |
| Pars opercularis | 1.024 (0.062) | 1.016 (0.060) | 0.005 | 0.429 | 0.652 |
| Rostral middle frontal | 1.024 (0.038) | 1.026 (0.044) | -0.003 | 0.436 | 0.652 |
| Superior frontal | 1.014 (0.043) | 1.017 (0.048) | -0.003 | 0.511 | 0.715 |
| Medial orbitofrontal | 1.012 (0.095) | 1.005 (0.085) | -0.004 | 0.520 | 0.717 |
| Precuneus | 1.009 (0.060) | 1.009 (0.056) | -0.003 | 0.587 | 0.771 |
| Postcentral | 1.025 (0.042) | 1.023 (0.044) | -0.002 | 0.636 | 0.808 |
| Parahippocampal | 0.997 (0.067) | 0.994 (0.068) | -0.003 | 0.659 | 0.814 |
| Posterior cingulate | 0.981 (0.050) | 0.981 (0.048) | -0.002 | 0.690 | 0.828 |
| Superior parietal | 1.051 (0.057) | 1.049 (0.051) | -0.002 | 0.713 | 0.843 |
| Pars triangularis | 1.024 (0.057) | 1.017 (0.062) | 0.002 | 0.749 | 0.850 |
| Pars orbitalis | 1.016 (0.048) | 1.012 (0.052) | -0.001 | 0.766 | 0.858 |
| Rostral anterior cingulate | 0.945 (0.056) | 0.943 (0.066) | 0.001 | 0.812 | 0.870 |
| Hippocampus | 0.964 (0.050) | 0.964 (0.051) | -0.001 | 0.818 | 0.870 |
| Inferior parietal | 1.032 (0.054) | 1.032 (0.052) | -0.001 | 0.832 | 0.873 |
| Substantia nigra | 0.920 (0.056) | 0.927 (0.054) | -0.001 | 0.889 | 0.911 |
| **Right hemisphere** |  |  |  |  |  |
| Precentral<sup>b</sup> | 1.016 (0.045) | 1.025 (0.048) | -0.015 | 0.002 | 0.090 |
| Supramarginal<sup>b</sup> | 1.000 (0.053) | 1.009 (0.050) | -0.014 | 0.007 | 0.146 |
| Superior temporal<sup>b</sup> | 0.994 (0.051) | 1.003 (0.047) | -0.014 | 0.007 | 0.146 |
| Cuneus<sup>b</sup> | 1.028 (0.080) | 1.032 (0.081) | -0.017 | 0.036 | 0.274 |
| Middle temporal<sup>b</sup> | 1.010 (0.062) | 1.017 (0.060) | -0.012 | 0.017 | 0.274 |
| Lingual<sup>b</sup> | 1.036 (0.064) | 1.039 (0.067) | -0.013 | 0.044 | 0.286 |
| Postcentral | 1.022 (0.051) | 1.029 (0.051) | -0.010 | 0.055 | 0.310 |
| Fusiform | 1.029 (0.088) | 1.030 (0.083) | -0.012 | 0.055 | 0.310 |
| Caudate | 1.083 (0.164) | 1.053 (0.144) | 0.025 | 0.107 | 0.428 |
| Parahippocampal | 1.003 (0.063) | 1.006 (0.057) | -0.008 | 0.133 | 0.429 |
| Inferior temporal | 1.056 (0.097) | 1.052 (0.090) | -0.009 | 0.150 | 0.435 |
| Amygdala | 1.002 (0.074) | 1.003 (0.068) | -0.009 | 0.147 | 0.435 |
| Rostral middle frontal | 1.015 (0.042) | 1.019 (0.049) | -0.006 | 0.211 | 0.452 |
| Superior frontal | 1.024 (0.048) | 1.028 (0.051) | -0.007 | 0.202 | 0.452 |
| Caudal middle frontal | 1.013 (0.066) | 1.022 (0.071) | -0.010 | 0.169 | 0.452 |
| Caudal anterior cingulate | 0.990 (0.079) | 0.978 (0.067) | 0.010 | 0.215 | 0.452 |
| Transverse temporal | 0.977 (0.069) | 0.981 (0.065) | -0.009 | 0.209 | 0.452 |
| Putamen | 0.920 (0.064) | 0.928 (0.059) | -0.008 | 0.164 | 0.452 |
| Pallidum | 0.914 (0.060) | 0.924 (0.057) | -0.007 | 0.204 | 0.452 |
| Hippocampus | 0.972 (0.051) | 0.976 (0.048) | -0.007 | 0.174 | 0.452 |
| Substantia nigra | 0.921 (0.055) | 0.931 (0.052) | -0.007 | 0.188 | 0.452 |
| Pericalcarine | 1.023 (0.091) | 1.024 (0.095) | -0.011 | 0.232 | 0.454 |
| Precuneus | 1.026 (0.064) | 1.027 (0.064) | -0.007 | 0.300 | 0.515 |
| Temporal pole | 1.022 (0.086) | 1.020 (0.077) | -0.008 | 0.283 | 0.515 |
| Banks STS | 0.989 (0.083) | 0.998 (0.072) | -0.008 | 0.297 | 0.515 |
| Lateral orbitofrontal | 1.002 (0.079) | 0.996 (0.075) | -0.006 | 0.337 | 0.566 |
| Lateral occipital | 1.035 (0.046) | 1.032 (0.051) | -0.004 | 0.385 | 0.614 |
| Medial orbitofrontal | 1.004 (0.087) | 0.999 (0.078) | -0.005 | 0.458 | 0.652 |
| Pars triangularis | 1.000 (0.066) | 1.003 (0.068) | -0.005 | 0.449 | 0.652 |
| Entorhinal | 1.068 (0.107) | 1.061 (0.100) | -0.006 | 0.456 | 0.652 |
| Isthmus cingulate | 0.989 (0.059) | 0.984 (0.060) | 0.004 | 0.542 | 0.734 |
| Inferior parietal | 1.033 (0.049) | 1.032 (0.049) | -0.003 | 0.580 | 0.771 |
| Insula | 0.973 (0.047) | 0.974 (0.042) | -0.002 | 0.602 | 0.778 |
| Pars orbitalis | 1.006 (0.054) | 1.001 (0.058) | -0.002 | 0.645 | 0.808 |
| Superior parietal | 1.049 (0.056) | 1.049 (0.050) | -0.002 | 0.674 | 0.821 |
| Pars opercularis | 1.008 (0.073) | 1.004 (0.066) | 0.003 | 0.725 | 0.846 |
| Frontal pole | 1.048 (0.056) | 1.041 (0.055) | 0.002 | 0.745 | 0.850 |
| Paracentral | 1.045 (0.087) | 1.040 (0.080) | -0.002 | 0.785 | 0.859 |
| Accumbens area | 1.008 (0.066) | 1.001 (0.060) | -0.001 | 0.787 | 0.859 |
| Posterior cingulate | 0.986 (0.052) | 0.981 (0.049) | 0.001 | 0.888 | 0.911 |
| Rostral anterior cingulate | 0.965 (0.080) | 0.962 (0.072) | 0.0004 | 0.957 | 0.969 |
| Thalamus | 0.992 (0.065) | 0.986 (0.062) | 0.00005 | 0.993 | 0.993 |

Regions are ordered separately for each hemisphere from the lowest FDR-adjusted $p$ -values to the highest.
<sup>a</sup>Significant when using the FDR-adjusted $p$ -values.
<sup>b</sup>Significant when using an uncorrected $p$ -value of $<0.05$ .
FDR = false discovery rate; iRBD = isolated REM sleep behaviour disorder; SD = standard deviation; STS = superior temporal sulcus.

**Supplementary Figure 1.**
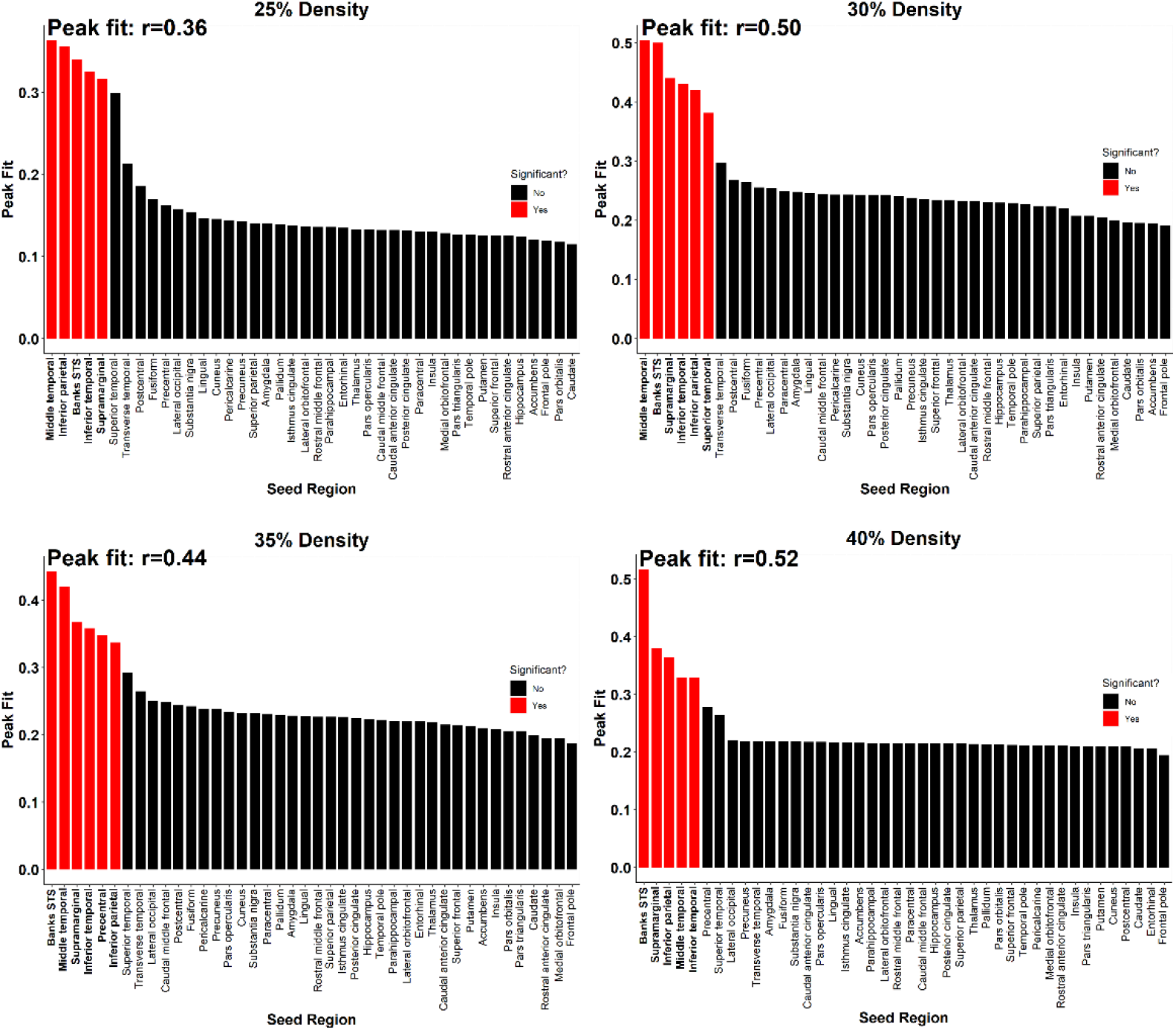
The model recreated deformation-based morphometry-derived tissue deformation of iRBD at different network densities. The atrophy simulated by the model using connection densities varying from 25% to 40% always recreated the pattern of tissue deformation observed in iRBD. Red columns represent the seed regions that led to a significant recreation of tissue deformation. iRBD = isolated REM sleep behaviour disorder; STS = superior temporal sulcus.

**Supplementary Figure 2.**
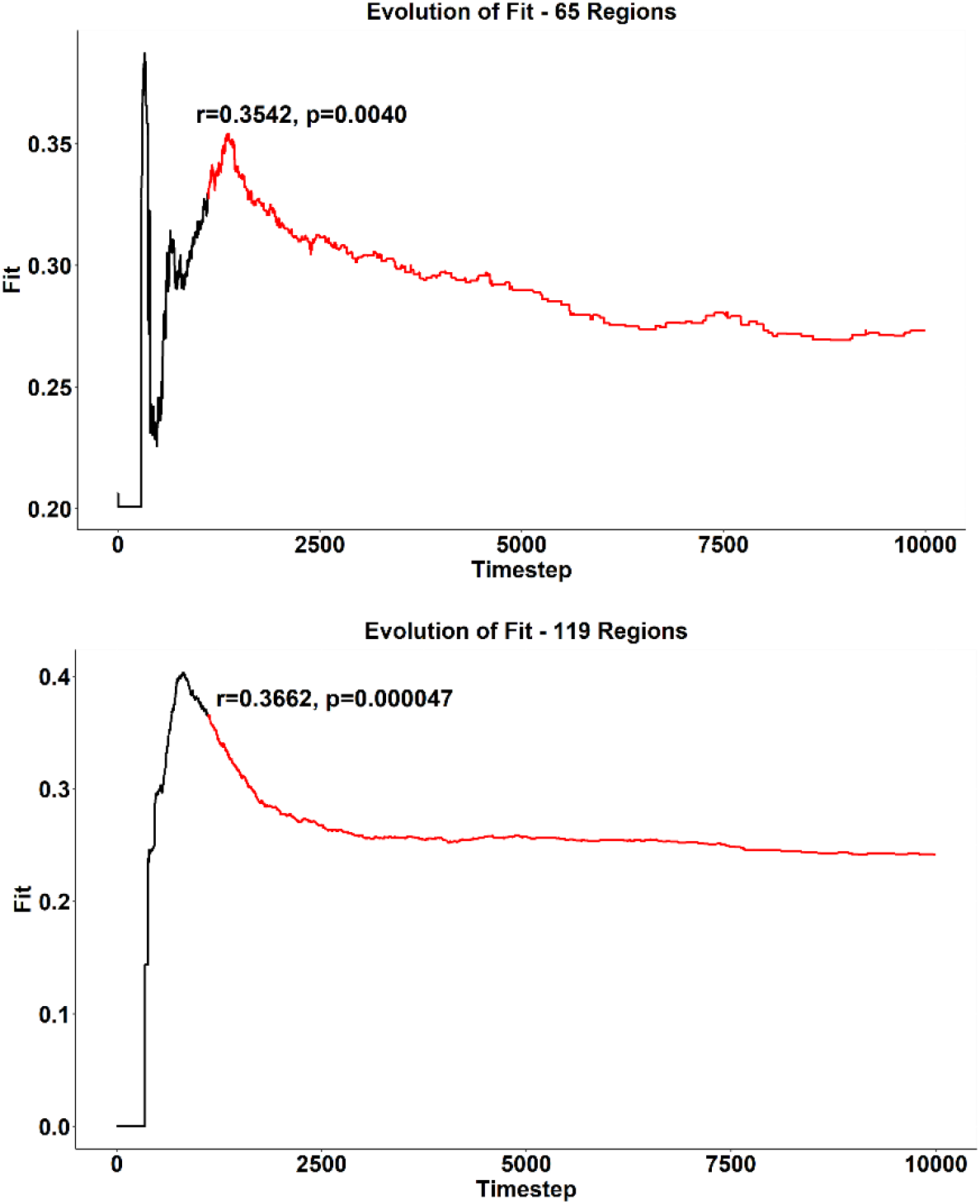
The model recreated the deformation-based morphometry-derived tissue deformation pattern using finer brain parcellations. The model also recreated the deformation-based morphometry-derived tissue deformation pattern of iRBD when simulating the spread of agents using brain parcellations of 65 and 119 regions. iRBD = isolated REM sleep behaviour disorder.

**Supplementary Figure 3.**
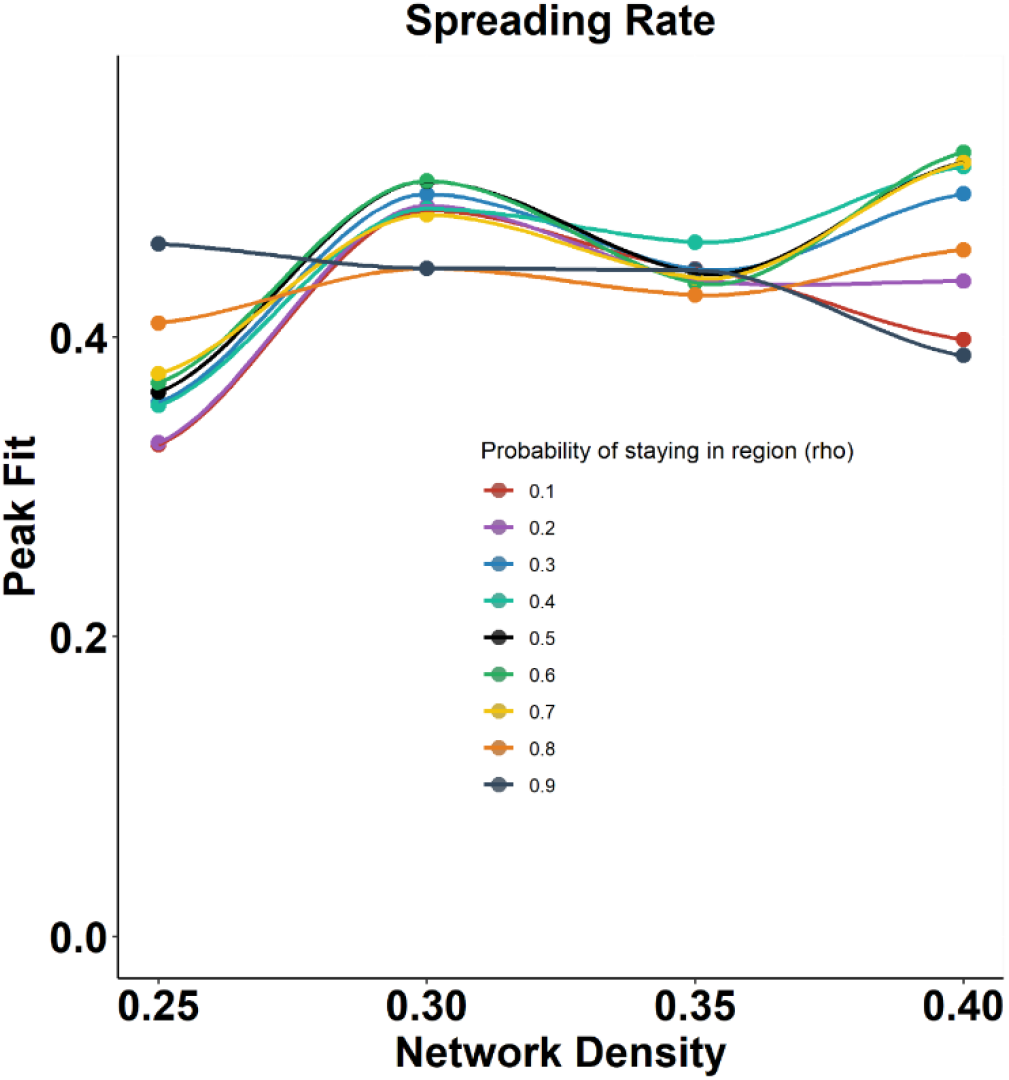
The model recreated the deformation-based morphometry-derived tissue deformation pattern at various spreading rates. The model recreated the deformation-based morphometry-derived tissue deformation pattern of iRBD when using different *ρ* values (i.e., the probability that an agent remained in a region instead of leaving the region) from 0.1 to 0.9. The main analyses performed in the article used *ρ*=0.5. iRBD = isolated REM sleep behaviour disorder.

**Supplementary Figure 4.**
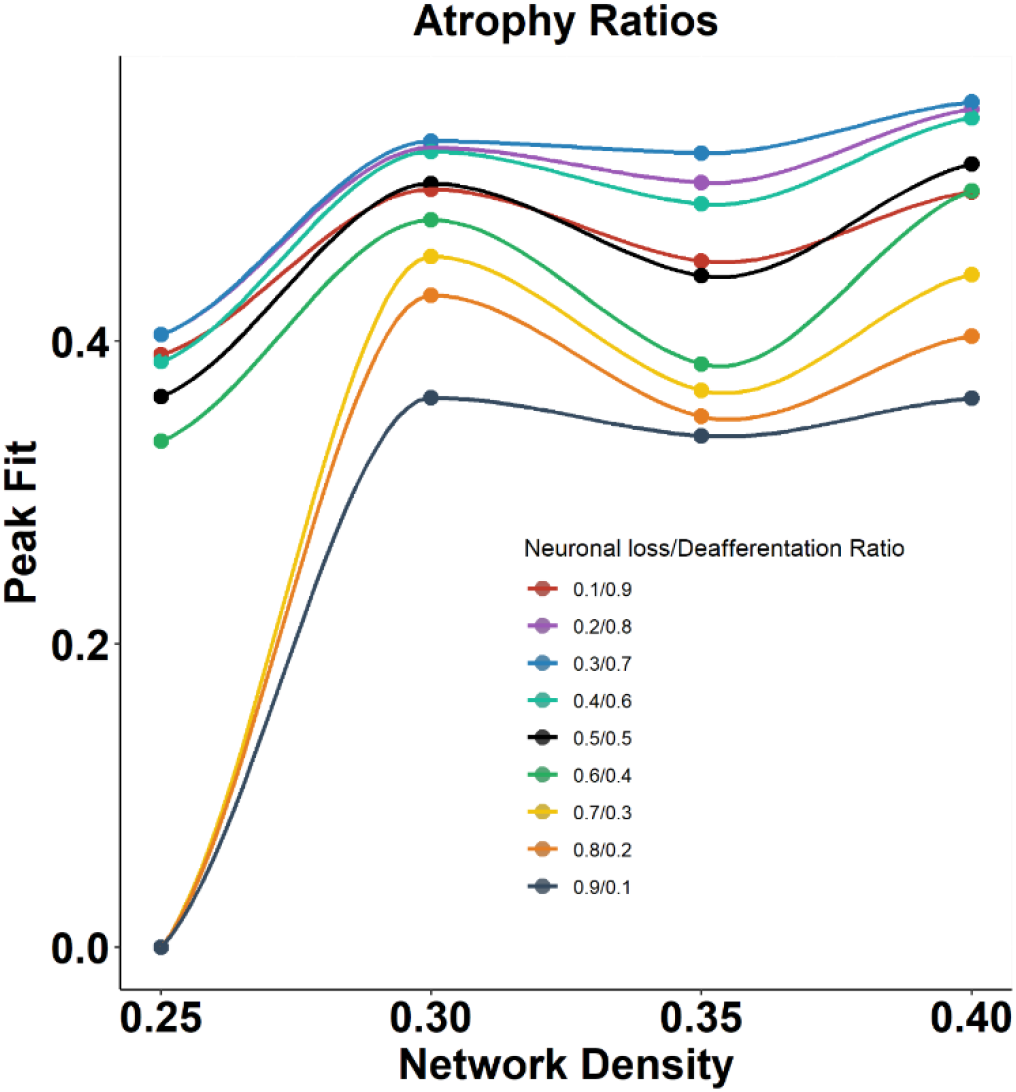
The model recreated the deformation-based morphometry-derived tissue deformation pattern at various atrophy ratios. The model recreated the deformation-based morphometry-derived tissue deformation pattern of iRBD when using different weights for neuronal loss and deafferentation in the quantification of the model’s simulated atrophy measure. The main analyses performed in the paper used equal weights for neuronal loss and deafferentation (*k*=0.5). iRBD = isolated REM sleep behaviour disorder.

